# Gamma Frequency Stimulation Provides Therapeutic Potential in Neuropsychiatry: A Systematic Review and Meta-Analysis

**DOI:** 10.64898/2026.04.10.26350641

**Authors:** Mei Xu, Angelina Singavarapu, Remi Philips, Monica Zheng, Donel Martin, Stevan Nikolin, Julian Mutz, Andrew Becker, Rosalind Firenze, Li-Huei Tsai

## Abstract

Gamma oscillation dysfunction has been implicated in neuropsychiatric disorders. Restoring gamma oscillations via brain stimulation represents an emerging therapeutic approach. However, the strength of its clinical effects and treatment moderators remain unclear. We conducted a systematic review and meta-analysis to examine the clinical effects of gamma frequency stimulation in neuropsychiatric disorders. A literature search for controlled trials using gamma frequency stimulation was performed across five databases up until April 2025. Effect sizes were calculated using Hedge’s g. Separate analyses using the random-effects model examined the clinical effects in schizophrenia (SZ), major depressive disorder (MDD), bipolar disorder, and autism spectrum disorder. For SZ and MDD, subgroup analyses evaluated the effects of stimulation modality, stimulation frequency, treatment duration, and pulses per session. Fifty-six studies met the inclusion criteria (N_SZ_ = 943, N_MDD_ = 916, N_BD_ = 175, N_ASD_ = 232). In SZ, gamma frequency stimulation was associated with improvements in positive (k = 10, g = −0.60, p < 0.001), negative (k = 12, g = −0.37, p = 0.03), depressive (k = 8, g = −0.39, p < 0.001), anxious symptoms (k = 5, g = −0.59, p < 0.001), and overall cognitive function (k = 7, g = 0.55, p < 0.001). Stimulation frequency and treatment duration moderated therapeutic effects. In MDD, reductions in depressive symptoms were observed (k = 23, g = −0.34, p = 0.007). However, confidence in these estimates is limited by substantial heterogeneity, and the predominance of 50 Hz theta-burst protocols.

## 1. Introduction

While gamma-band dysfunction, i.e., the disruption of synchronized neuronal activity in the 30–100 Hz range, represents an emerging biomarker in psychiatric care, its potential as a therapeutic target has not been fully explored. Traditional treatments rarely target physiological disturbances in fast-frequency neuronal oscillations, which have been associated with higher order cognitive functions. Antipsychotics poorly address cognitive deficits; antidepressants do not restore aberrant cortical synchrony; psychosocial treatments rely on intact neurocognitive systems that may themselves be impaired (Strauss and Cohen, 2017). Here, we systematically evaluate the existing literature on the effectiveness of brain stimulation in the gamma range as a safe and scalable approach to directly modulate gamma dysfunctional neural activity and restore network-level abnormalities.

### 1.1. Gamma Oscillation

Gamma oscillations, detectable by electroencephalography (EEG), comprise at least two partially distinct rhythms, slow gamma (30–50 Hz) and fast gamma (50–100 Hz), arising from local circuit interactions involving overlapping populations of GABAergic interneurons (Guan et al., 2022; Mably and Colgin, 2018). Slow gamma, in particular, supports perceptual integration, working memory, attentional control, and large-scale neural communication (Strüber and Herrmann, 2020). Abnormalities in this gamma band are closely associated with cognitive impairment, sensory integration deficits, and affective dysregulation (Herrmann and Demiralp, 2005), positioning gamma dysfunction as a mechanistic contributor to circuit-level dyscoordination (Hanslmayr et al., 2019). Acute and chronic intracranial EEG recordings show hippocampal slow gamma frequency strength to predict associative memory (Henin et al., 2019) and verbal memory (Sederberg et al., 2007).

### 1.2. Gamma Dysfunction in Neuropsychiatric Conditions

Gamma oscillation impairments, particularly in the slow gamma range, are widely documented across neuropsychiatric and neurodegenerative disorders, including schizophrenia (SZ), bipolar disorder (BD), attention deficit hyperactivity disorder (ADHD), autism spectrum disorder (ASD) and alzheimer’s disease (AD) (Deng et al., 2024), pointing to shared neurophysiological dysfunctions and motivating a growing interest in gamma-targeted interventions. Disruption of gamma activity is particularly well characterized in SZ, where reduced gamma power and impaired phase synchronization are consistently observed, especially in auditory steady-state responses (ASSRs) (Kinjo et al., 2026; Thuné et al., 2016). The 40 Hz ASSR has emerged as a transdiagnostic marker of cognitive dysfunction across SZ (Thuné et al., 2016), AD (Shahmiri et al., 2017) and BD (Jefsen et al., 2022), with reduced gamma synchrony associated with working memory impairment and broader cognitive deficits (Koshiyama et al., 2021). Converging evidence implicates reduced inhibitory control due to dysfunction of GABAergic and, in particular, parvalbumin-positive (PV) interneurons and N-methyl-D-aspartate (NMDA) receptor hypofunction in these impairments (Chen et al., 2014; Lewis, 2014; Lewis et al., 2012).

### 1.3. Gamma Frequency Stimulation in Neuropsychiatric Conditions

Recent studies demonstrate the therapeutic potential of gamma frequency neuromodulation by invasive and non-invasive brain stimulation in AD, Parkinson’s diseases (PD), SZ, mood disorders, and neurodevelopmental disorders. Invasive brain stimulation (e.g., deep brain stimulation) mostly used high gamma frequency (50–100 Hz). Non-invasive neuromodulation commonly using a lower gamma frequency range (30–50 Hz) can be achieved through transcranial magnetic stimulation (TMS), sensory stimulation, transcranial alternating current stimulation (tACS), and vagus nerve stimulation (VNS) (Fig. 1).

**Fig. 1.**
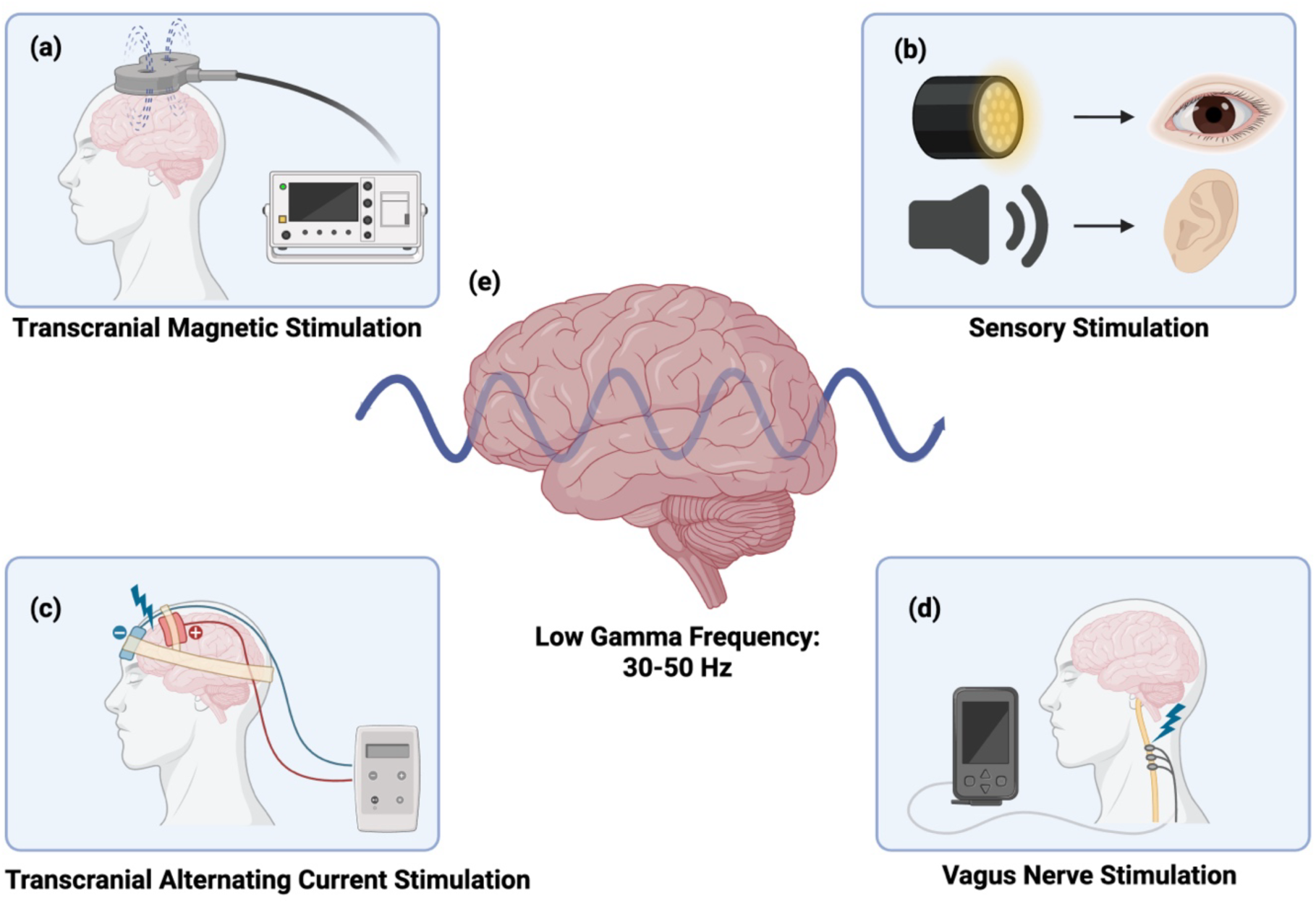
Non-invasive Gamma Frequency Stimulation. Note: Different neuromodulation modalities are utilized to deliver gamma frequency stimulation. (a) Transcranial Magnetic Stimulation (TMS), following Faraday’s principle, delivers a magnetic field to non-invasively stimulate cortical brain regions at the gamma frequency range. (b) Sensory stimulation (SS) using light and sound stimuli is transduced by sensory organs, and the signal propagates through sensory processing pathways to target regions. (c) Transcranial alternating current stimulation (tACS) passes alternating electrical current between scalp electrodes to entrain neural activity at specific frequencies, including gamma. (d) Vagal nerve stimulation (VNS) leverages the vagus nerve’s widespread projections to modulate gamma-frequency activity across distributed targets. (e) Non-invasive gamma stimulation for therapeutic purposes typically uses low gamma frequencies between 30 Hz and 50Hz. This figure was created with Biorender.

Repetitive TMS (rTMS, Fig. 1a), based on the principles of electromagnetic induction to trigger neuronal firing, is the most widely used technique (Barker et al., 1985). Theta-burst stimulation (TBS) represents a patterned form of rTMS with 50 Hz bursts delivered at a theta frequency of 5 Hz (Huang et al., 2005), either continuously (cTBS) to produces inhibitory aftereffects or intermittently (iTBS), i.e., separated by an 8s inter-train interval, to induce excitatory aftereffects (Huang et al., 2005). 40 Hz repetitive TMS and TBS have demonstrated clinical potential in SZ (Kishi et al., 2024a), mood disorders (Kishi et al., 2024b), and neurodevelopmental disorders (Yuan et al., 2024). Sensory stimulation, which is based on auditory and/or visual stimulation at gamma frequencies (Fig. 1b), can reliably entrain gamma oscillations and induce cognitive benefits (Hajós et al., 2024), with therapeutic potential reported in neurodegenerative and mood disorders including AD and MDD [24,25]. Particularly at 40 Hz (Chan et al., 2022, 2021), gamma entrainment using sensory stimuli (GENUS) has shown promise in enhancing neural synchrony and mitigating cognitive decline, although the number of studies remains limited [26, 27]. Gamma-frequency tACS (e.g., 30–40 Hz) uses electrical stimulation (see Fig. 1c), to enhance synaptic plasticity and cognitive function by synchronizing cortical networks (Sánchez-Garrido Campos et al., 2025), while non-invasive VNS (see Fig. 1d) delivers pulsed electrical stimulation to modulate brain-body circuits implicated in mood regulation (Garcia et al., 2021). Although these interventions show potential for improving cognitive and clinical outcomes in neuropsychiatric populations (Alexander et al., 2019a; Cao et al., 2025; Garcia et al., 2021), current findings for electrical stimulation modalities are mixed.

### 1.4. Critical Moderators of Gamma Frequency Stimulation

The effects of gamma frequency modulation critically depend on treatment parameters, including stimulation modality, stimulation frequency, and dose per session. For example, iTBS and cTBS using 50 Hz magnetic stimulation generate the opposite effects on the motor cortex (Huang et al., 2005). Similarly, sensory stimulation increased the cognitive performance (Chan et al., 2025, 2022), and alleviated depressive symptoms within the 30–50 Hz range (Lee et al., 2021). Other important moderators include treatment duration (e.g., 1-4 weeks) and total pulses per session (e.g., 600 pulses, 1800 pulses) (Holczer et al., 2020). However, it remains unclear how stimulation parameters contribute to the therapeutic effects of gamma frequency stimulation across different neuropsychiatric illnesses.

A rigorous synthesis of the existing evidence is therefore needed to determine whether gamma-targeted interventions constitute a viable, mechanism-informed approach for improving clinical and cognitive outcomes. In this review, we quantitatively assess the clinical efficacy and cognitive effects of gamma frequency stimulation in neuropsychiatric disorders and examine key moderators of treatment response. Our hypotheses include: (1) active gamma frequency neuromodulation has a superior effect on clinical symptoms in neuropsychiatric disorders compared to the control condition; (2) active gamma frequency neuromodulation enhances cognitive functions in neuropsychiatric disorders compared to the control condition; (3) stimulation modalities, stimulation frequency, stimulation duration, and total pulse per session are critical factors in moderating treatment effects.

## 2. Methods

Our systematic review and meta-analysis followed the guidelines outlined in the Cochrane Handbook for Systematic Reviews of Interventions (Higgins and Green, 2008) and the PRISMA guidelines (Liberati et al., 2009). The search protocol for this study was registered on the PROSPERO international prospective protocol for systematic reviews (PROSPERO 2025 CRD420251030517).

### 2.1. Literature search

A literature search was conducted by three authors (MX, RP, and MZ) across PubMed, MEDLINE, Embase, PsycINFO, and Web of Science from their inception through April 21, 2025. Inclusion criteria were: (1) psychiatric patients with evidence of abnormalities or disruption in gamma oscillations in the brain, including ASD, ADHD, SZ, BD, and MDD; (2) gamma (30–50 Hz) frequency neuromodulation including TMS, deep TMS, tACS, audiovisual entrainment, sensory stimulation, GENUS, VNS, deep brain stimulation, ultrasound stimulation, subcutaneous electrical stimulation; (3) active control or sham treatment groups; (4) assessments of clinical evaluation and cognitive tasks; (5) Controlled trials with either parallel or cross-over designs were included, while case studies were excluded. Only human studies published in English were considered. The search strategy is summarised in the supplementary material.

### 2.2. Study selection

Review articles, conference abstracts, and duplicate records were removed using Covidence (Veritas Health Innovation, 2025). Studies were then assessed against the predefined inclusion criteria. Two authors (RP and MZ) independently screened titles, abstracts, and full-text articles identified in the search (see Fig. S1). Any disagreements regarding study eligibility were resolved through discussion with the senior investigator (MX).

### 2.3. Risk of bias assessment

The risk of bias across included studies was independently evaluated by two reviewers (RP and MZ) using the RoB 2 tool (Sterne et al., 2019). All included studies were evaluated across the following domains: (1) bias arising from the randomization process; (2) bias due to deviations from intended interventions; (3) bias resulting from missing outcome data; (4) bias in outcome measurement; and (5) bias in the selection of reported results. Each domain was rated as “low risk of bias,” “some concerns,” or “high risk of bias.” Any disagreements were resolved through discussion with the lead investigator (MX).

### 2.4. Data extraction

Data extraction was carried out by MX and covered clinical and cognitive outcomes alongside supplementary study information, including active and control group sample sizes, participant demographics (age, gender, and education level), treatment parameters (frequency, number and duration of sessions, and pulses per session), clinical and cognitive assessments, and post-gamma stimulation means and standard deviations (SD) or standard mean differences (SEM). In cases where data appeared only in figures, values were retrieved with the aid of WebPlotDigitizer 5.2 (Rohatgi, 2020). When needed, we reached out to the first or corresponding authors of included studies to request data or further information.

### 2.5. Quantitative synthesis

From the studies identified, we gathered clinical assessment outcomes following stimulation for both the active and control groups. Clinical assessments were categorized based on clinical symptoms under four different disorders (SZ, MDD, BD, and ASD). Based on post-stimulation scores across active and control conditions, effect sizes were computed (Higgins et al., 2019). When the number of studies for a given symptom or subgroup fell below five, this was regarded as too limited for inclusion in a quantitative meta-analysis. (Jackson and Turner, 2017). Hedge’s g was used to estimate effect sizes for both clinical and cognitive datasets, expressed as the standardized mean difference at post-treatment (Higgins and Green, 2008). Where multiple outcomes were available for a single task within a study, we prioritized the effect size of the primary outcome measure as specified by the original authors (Begemann et al., 2020). When no primary outcome measure was specified for clinical assessments or cognitive tasks, the most relevant measure was selected based on the disorder-specific criteria outlined in Table 1.

**Table 1.**
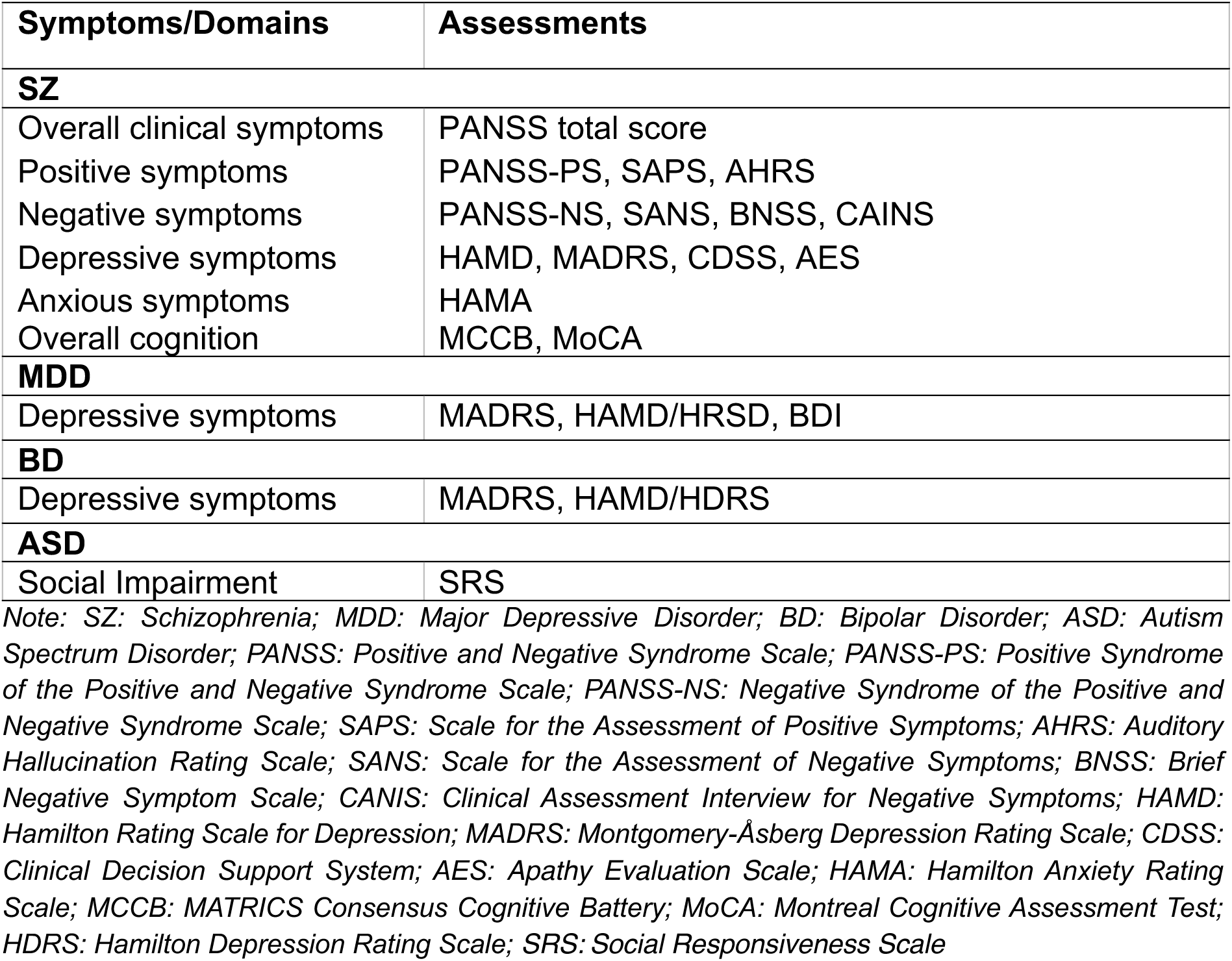
Summary of outcome measures and included clinical and cognitive assessments.

All statistical analyses were performed in R (version 4.3.0) (R Core Team, 2023) and RStudio (version 2023.03.0+386) (Posit team, 2022), utilizing the ‘*meta*’ and ‘*metafor*’ packages. Clinical and cognitive outcomes were each examined through separate meta-analyses employing random-effects models with the Paule and Mandel (PM) estimator (Veroniki et al., 2016). Cochran’s I² statistic was used to assess heterogeneity, while small-study effects were evaluated through visual inspection of funnel plots and Egger’s test (Egger et al., 1997). Sensitivity analyses were carried out by excluding studies identified as outliers, meaning those whose confidence intervals showed no overlap with that of the pooled effect estimate (Viechtbauer and Cheung, 2010). Additional sensitivity analyses were also conducted by removing cTBS or iTBS studies from overall analyses. To obtain sufficient statistical power to detect bias, Egger’s test was performed when the number of studies was more than 10. To detect and account for possible publication bias, we applied the trim and fill method (Duval and Tweedie, 2000). For each of the four disorders, clinical assessments and cognitive tasks were examined in independent meta-analyses. Additional subgroup analyses were carried out to explore the effects of stimulation modality, frequency, treatment duration, and total pulses per session.

## 3. Results

### 3.1. Search results

A total of 60 studies met our inclusion criteria. Four publications lacked sufficient data for extraction (Baeken et al., 2019, 2017; Cheng et al., 2023; DeMayo et al., 2024), resulting in 56 studies being incorporated into the quantitative synthesis (Alexander et al., 2019a; Basavaraju et al., 2021; Batail et al., 2023; Bation et al., 2021; Bodén et al., 2021; Cao et al., 2025; Chauhan et al., 2021; Chen et al., 2019a; Chou et al., 2023; Dellink et al., 2024; Diederichs et al., 2021; Duprat et al., 2016; Fitzgerald, 2021; Gao et al., 2025; Garcia et al., 2021; Garg et al., 2022; Holczer et al., 2021; Hoy et al., 2016; Hua et al., 2024; Huang et al., 2025; Ji et al., 2023; Jin et al., 2023; Kang et al., 2024; Koops et al., 2016; Kos et al., 2024; Li et al., 2014a, 2014b, 2023; J. Li et al., 2025; L. Li et al., 2025, 2025; Li et al., 2024; Liu et al., 2025; Luo et al., 2024; Mallik et al., 2023; McGirr et al., 2021a; Ni et al., 2017, 2021, 2022, 2023, 2024; Noda et al., 2025; Sheline et al., 2024; Shinn et al., 2023, 2023; Stöhrmann et al., 2023; Tavares et al., 2021, 2021; Tikka et al., 2017; Tyagi et al., 2022; Vergallito et al., 2024; Walther et al., 2020, 2020; Wang et al., 2020, 2022; Wilkening et al., 2022; Wu et al., 2021; Zavorotnyy et al., 2020; Zhang et al., 2024; Zhao et al., 2023; Zhu et al., 2021). The search and screening workflow is illustrated in Fig.S1.

### 3.2. Risk of bias results

Overall, the risk of bias was performed across 56 studies. 10% of studies were rated as having high risk of bias, 62% had some concerns, and 28% were judged to have low risk (Fig. S2). Most studies demonstrated low risk in the domains of outcome measurement (88%), missing outcome data (77%), deviations from intended interventions (70%), and randomization procedures (75%). In terms of reported results selection, 58% of included studies were deemed low risk, with the remaining studies flagging concerns related to the absence of preregistered protocols or predefined analysis plans.

### 3.3. Study characteristics

Table S1 and Fig. 2 present the characteristics of the included studies. Most investigations were conducted in Asia, followed by North America, Australia, and South America. Nearly half of the studies (48%) examined gamma frequency stimulation in SZ, with additional studies in MDD (35%), BD (9%), and ASD (9%). The current analyses focused on non-invasive brain stimulation using 30 Hz to 50 Hz. ITBS was the predominant modality (64%), followed by cTBS (21%), tACS (7%), bilateral TBS (5%), sensory stimulation (2%), and VNS (2%). Stimulation frequencies were 30 Hz (3%), 40 Hz (9%), and 50 Hz (87.9%), which was most commonly employed in magnetic-stimulation protocols. Intervention duration varied, from a 2-week schedule adopted by 34.5% of studies and 1-week (21%), 3-week (19%), and 4-week (17%) protocols used at similar rates. Session doses varied between 1500–2000 and 1000–1500 pulses per session in 35% and 16%, respectively, of studies and both 500–1000 and >2000 pulses in approximately one-quarter of studies.

**Fig. 2.**
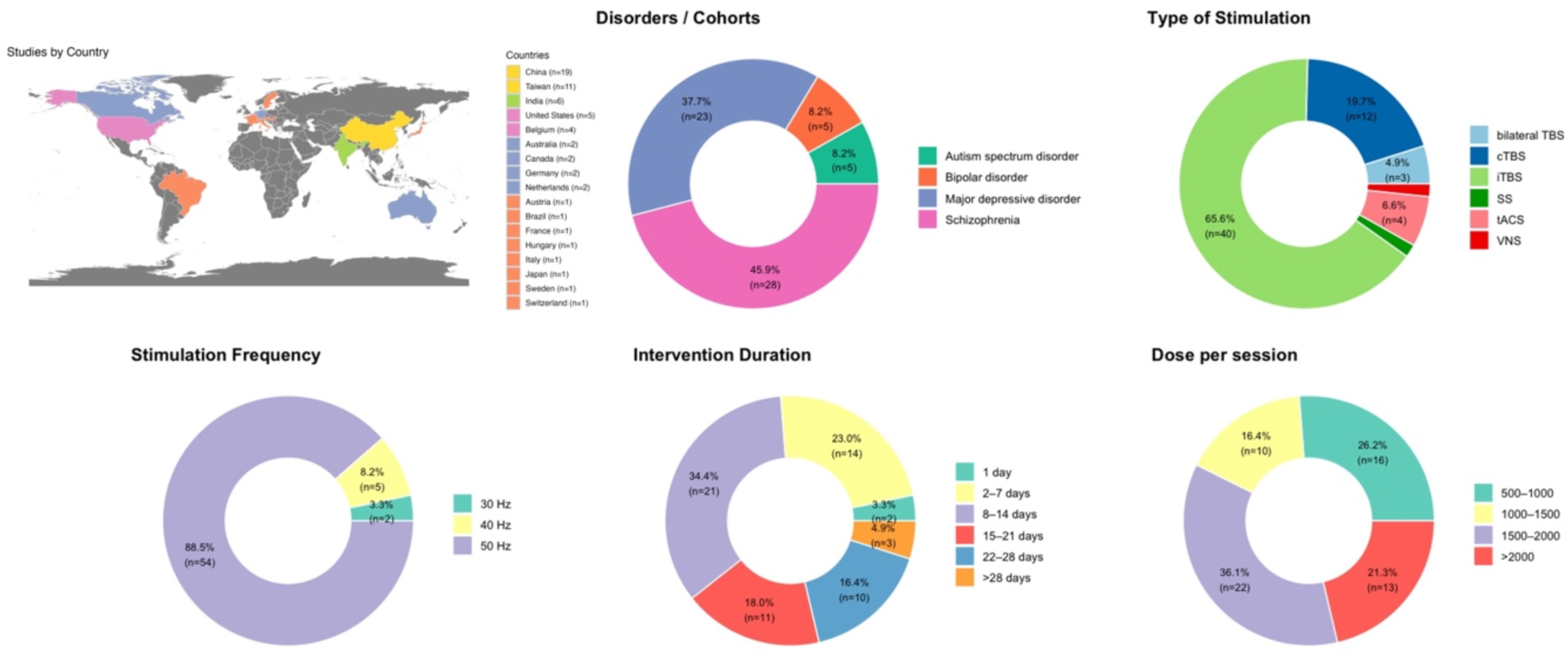
Study Characteristics Summary. Note: tACS: Transcranial Alternating Current Stimulation; cTBS: Continuous Theta Burst Stimulation; iTBS: Intermittent Theta Burst Stimulation; BiTBS: Bilateral Theta Burst Stimulation; VNS: Vagus Nerve Stimulation; SS: Sensory Stimulation.

### 3.4. Clinical symptoms and cognition

Table 2 summarizes the effect sizes of non-invasive gamma frequency stimulation on clinical symptoms and cognitive functions across SZ, MDD, BD, and ASD. Overall, non-invasive gamma frequency stimulation modulated clinical symptoms in SZ and MDD and enhanced cognitive functions in SZ compared to controls.

**Table 2.**
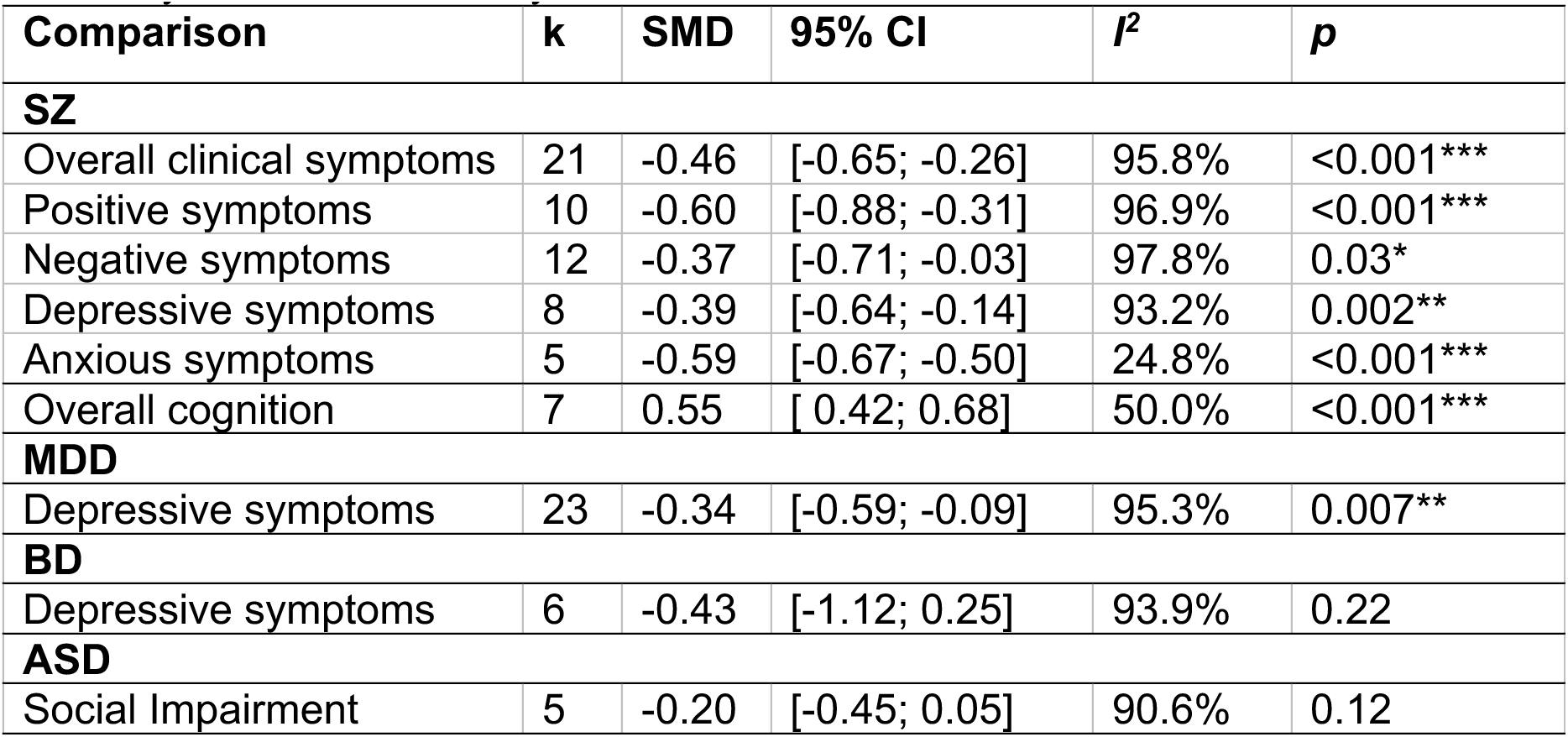
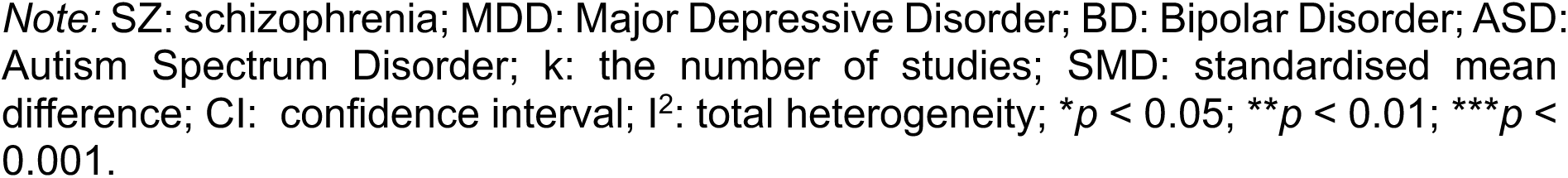
Summary table of meta-analysis results across disorders.

| Comparison | k | SMD | 95% CI | I<sup>2</sup> | p |
| --- | --- | --- | --- | --- | --- |
| **SZ** |  |  |  |  |  |
| Overall clinical symptoms | 21 | -0.46 | [-0.65; -0.26] | 95.8% | <0.001*** |
| Positive symptoms | 10 | -0.60 | [-0.88; -0.31] | 96.9% | <0.001*** |
| Negative symptoms | 12 | -0.37 | [-0.71; -0.03] | 97.8% | 0.03* |
| Depressive symptoms | 8 | -0.39 | [-0.64; -0.14] | 93.2% | 0.002** |
| Anxious symptoms | 5 | -0.59 | [-0.67; -0.50] | 24.8% | <0.001*** |
| Overall cognition | 7 | 0.55 | [ 0.42; 0.68] | 50.0% | <0.001*** |
| **MDD** |  |  |  |  |  |
| Depressive symptoms | 23 | -0.34 | [-0.59; -0.09] | 95.3% | 0.007** |
| **BD** |  |  |  |  |  |
| Depressive symptoms | 6 | -0.43 | [-1.12; 0.25] | 93.9% | 0.22 |
| **ASD** |  |  |  |  |  |
| Social Impairment | 5 | -0.20 | [-0.45; 0.05] | 90.6% | 0.12 |

Note: SZ: schizophrenia; MDD: Major Depressive Disorder; BD: Bipolar Disorder; ASD: Autism Spectrum Disorder; k: the number of studies; SMD: standardised mean difference; CI: confidence interval; $I^2$ : total heterogeneity; \* $p < 0.05$ ; \*\* $p < 0.01$ ; \*\*\* $p < 0.001$ .

#### 3.4.1. Schizophrenia (SZ)

The overall effect of active gamma frequency stimulation (30 Hz, 40 Hz, 50 Hz) using total Positive and Negative Syndrome Scale (PANSS) scores was statistically significant in SZ compared to the control groups (k = 21, g = -0.46, 95% CI = [-0.65; - 0.26], p < 0.001, Fig. 3a and Table 2), with substantial heterogeneity (I^2^ = 96.0%). Following removal of eight outlier studies, the effect of active gamma frequency stimulation remained statistically significant compared to control (k = 13, g = -0.46, 95% CI = [-0.59; -0.33], p < 0.001), with heterogeneity remaining high (I^2^ = 80.2%). The contoured funnel plot suggested some possibility of publication bias upon visual inspection; however, this was not supported by the Egger’s test results (p = 0.32, see Fig. S3). In sensitivity analyses, the overall effect stayed at -0.39 (k = 15, 95% CI = [-0.62; -0.16], p < 0.001) after removing cTBS studies.

**Fig. 3.**
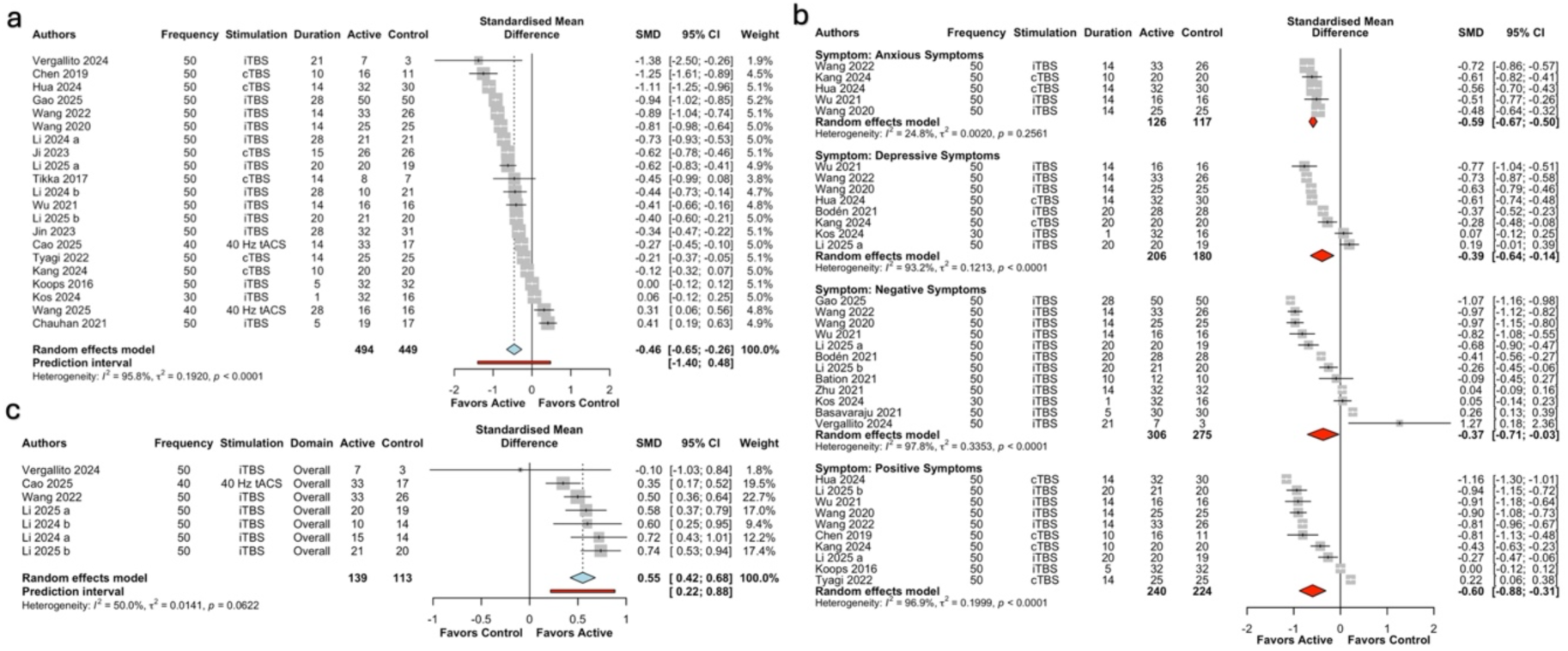
Forest plot of gamma frequency stimulation in SZ. *Note:* Duration: treatment period (days). iTBS: Intermittent Theta Burst Stimulation; tACS: Transcranial Alternating Current Stimulation; cTBS: Continuous Theta Burst Stimulation; SMD: Standardised Mean Difference; I^2^: total heterogeneity; \**p* < 0.5; \*\**p* < 0.01; \*\*\**p* < 0.001.

##### Positive, negative, depressive, and anxious symptoms

Compared to controls, active gamma frequency stimulation (50 Hz) had a significant moderate-sized effect on positive symptoms (k = 10, g = -0.60, 95% CI = [-0.88; -0.31], p < 0.001, I^2^ = 96.9%); a statistically significant medium-sized effect on negative symptoms (k = 12, g = -0.37, 95% CI = [-0.71; -0.03], p = 0.03, I^2^ = 97.8%); a statistically significant small-sized effect on executive function (k = 8, g = -0.39, 95% CI = [-0.64; -0.14], p = 0.002, I^2^ = 93.2%); and a statistically significant medium-sized effect on anxious symptoms (k = 5, g = -0.59, 95% CI = [-0.67; -0.50], p < 0.001, I^2^ = 24.8%) (Fig. 3b and Table 2). No publication bias was detected by the Egger’s test for positive (p = 0.40) or negative (p = 0.41) symptoms.

##### Cognitive function

Active gamma frequency stimulation (40 Hz: k = 1, 50 Hz: k = 7), including tACS and iTBS, enhanced global cognitive functioning on the MATRICS Consensus Cognitive Battery and Montreal Cognitive Assessment Test (k = 7, g = 0.55, 95% CI = [0.42; 0.68], p < 0.001) compared to controls with heterogeneity at 50.0% (Fig. 3c and Table 2). Other cognitive domains had fewer than five studies investigating the cognitive effect of gamma frequency stimulation; no further meta-analyses were conducted.

##### Subgroup analyses

For the overall effect, subgroup analyses of active gamma frequency stimulation (Fig. 4 and Table S2) revealed a statistically significant effect of stimulation frequency (30 Hz: k = 1, g = 0.06; 40 Hz: k = 2, g = 0.01; 50 Hz: k = 18, g = -0.54; p < 0.001) and treatment duration (<1 week: k = 1, g = 0.06; 1-2 week: k = 4, g = -0.23; 2-3 week: k = 8, g = -0.61; >3 week: k = 8, g = -0.50; p < 0.001). Among the tested parameters, moderate effect sizes (g ≥ 0.5) were indicated for a stimulation frequency of 50 Hz and treatment durations of 2-3 weeks or more. No significant differences were found for stimulation modalities and pulses per session.

**Fig. 4.**
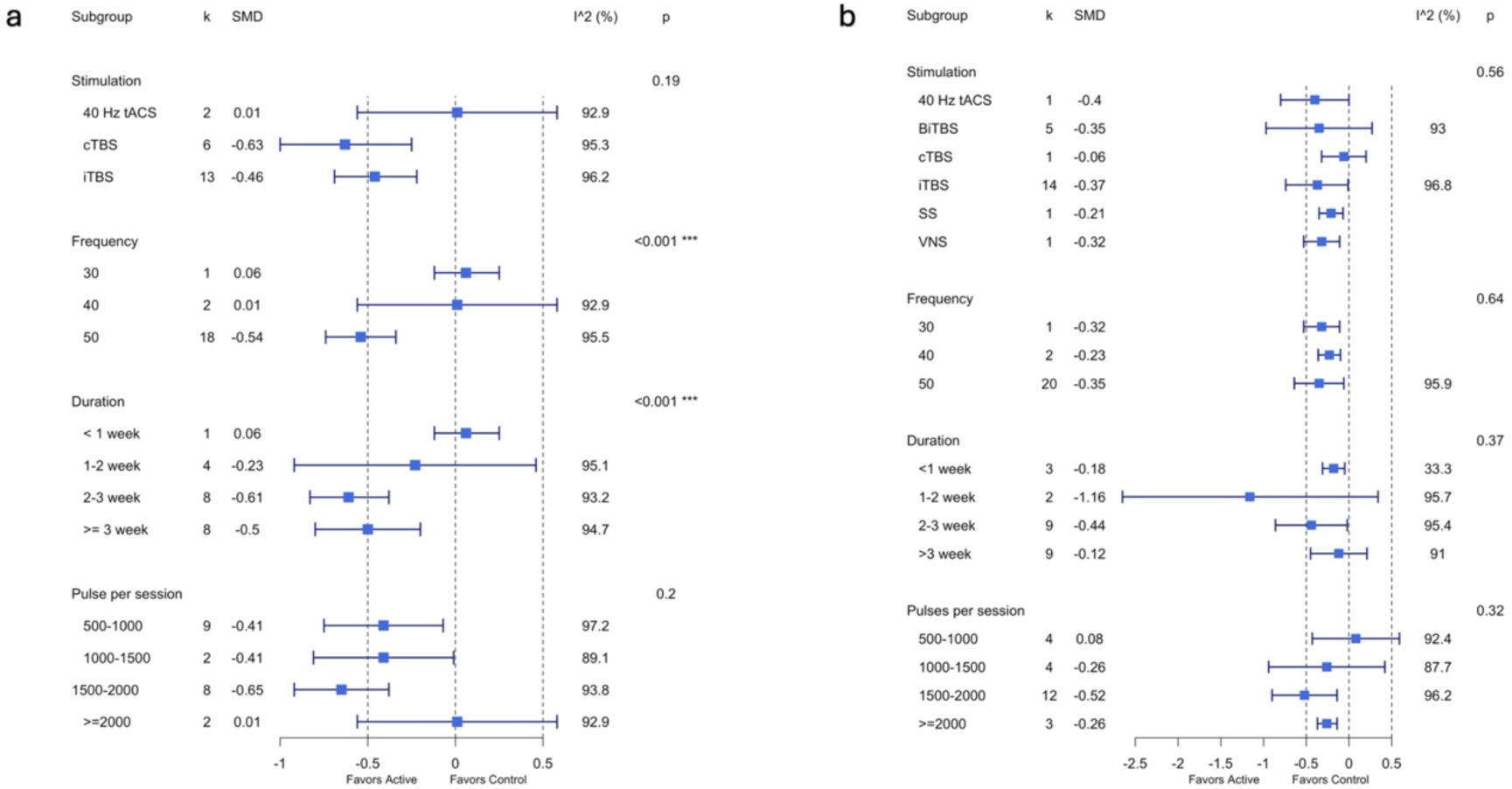
Subgroup-analysis plot in SZ and MDD. *Note:* tACS: Transcranial Alternating Current Stimulation; cTBS: Continuous Theta Burst Stimulation; iTBS: Intermittent Theta Burst Stimulation; BiTBS: Bilateral Theta Burst Stimulation; VNS: Vagus Nerve Stimulation; SS: Sensory Stimulation; SMD: Standardised Mean Difference; I^2^: total heterogeneity; \**p* < 0.5; \*\**p* < 0.01; \*\*\**p* < 0.001.

#### 3.4.2. Major depressive disorder (MDD)

##### Depressive and anxious symptoms

Active gamma frequency stimulation at 30, 40, or 50 Hz demonstrated a statistically significant overall effect on depressive symptoms compared with control conditions (k = 23, g = -0.34, 95% CI = [-0.59; -0.09], p < 0.001), accompanied by substantial heterogeneity (I² = 95.3%) (Fig. 5 and Table 2). The effect remained statistically significant and of small magnitude after excluding nine statistical outliers (k = 14, g = -0.30, 95% CI = [-0.42, -0.19], p < 0.001), with heterogeneity persisting at a high level (I^2^ = 76.9%). In sensitivity analyses, after removing cTBS studies and both iTBS and cTBS studies, the effect size remained at -0.36 (k = 22, 95% CI = [-0.62; -0.09], p < 0.01) and -0.26 (k = 3, 95% CI = [-0.42, -0.19], p < 0.001) respectively. Visual inspection of the funnel plot and the Egger’s test (p = 0.004) revealed risk of publication bias (Fig. S4). The trim and fill method was used to adjust the funnel plot asymmetry, then the publication bias disappeared. As there were fewer than five studies investigating the effect of gamma frequency stimulation on anxious symptoms, meta-analyses were not conducted.

**Fig. 5.**
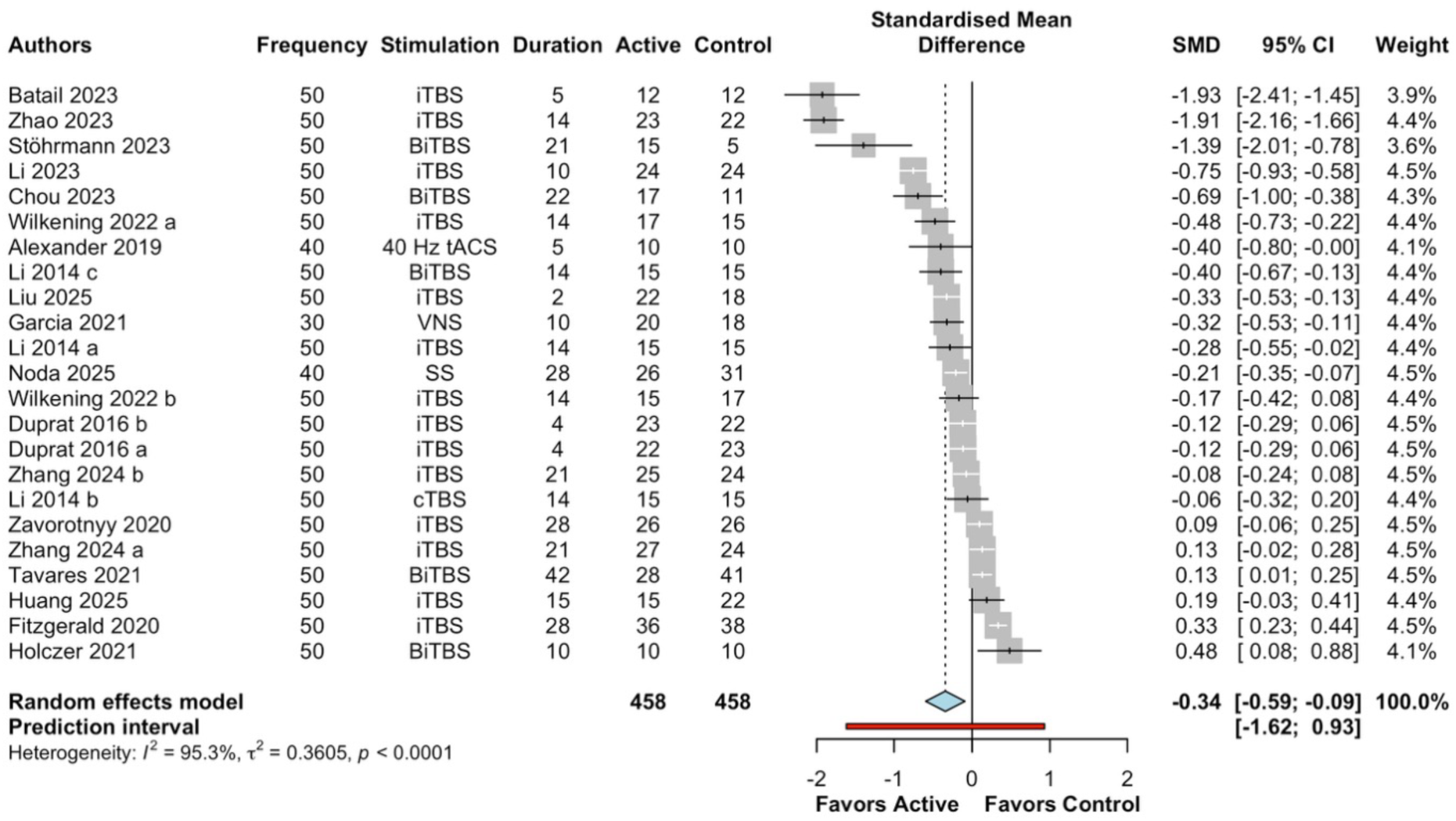
Forest plot of gamma frequency stimulation on depressive symptoms in MDD. *Note:* Duration: treatment period (days). iTBS: Intermittent Theta Burst Stimulation; BiTBS: Bilateral Theta Burst Stimulation; tACS: Transcranial Alternating Current Stimulation; VNS: Vagus Nerve Stimulation; SS: Sensory Stimulation; cTBS: Continuous Theta Burst Stimulation; SMD: Standardised Mean Difference; I^2^: total heterogeneity; \**p* < 0.5; \*\**p* < 0.01; \*\*\**p* < 0.001.

##### Subgroup analyses

No significant finding of stimulation modality, stimulation frequency, duration, or total pulses per session was reported for moderating depressive symptoms (Fig.3 and Table S2).

#### 3.4.3. Bipolar disorder (BD)

##### Depressive, anxious, and manic symptoms

Relative to control conditions, active gamma frequency stimulation (50 Hz) had a positive effect on depressive symptoms, but the effect was not statistically significant (k = 6, g = -0.43, 95% CI = [-1.12; 0.25], p = 0.22), with heterogeneity at 93.9% (Table 2 and Fig. S5). Given that fewer than five studies examined the effects of gamma frequency stimulation on anxiety or manic symptoms (Table 2; Fig. S5), meta-analyses were not performed.

#### 3.4.4. Autism spectrum disorder (ASD)

##### Social impairment, other clinical symptoms, and cognitive function

Active gamma frequency stimulation (50 Hz) had a positive effect on social impairment in comparison with controls, but the effect was not statistically significant (k = 5, g = - 0.20, 95% CI = [-0.45; 0.05], p = 0.12), with heterogeneity at 90.6% (Table 2 and Fig. S6). As there were fewer than five studies investigating the effect of gamma frequency stimulation on other ASD relevant symptoms and cognitive function (Table 2, Fig. S6 and S7), meta-analyses were not conducted.

## 4. Discussion

To the best of our knowledge, no prior systematic review or meta-analysis has examined the therapeutic efficacy of gamma-frequency stimulation across neuropsychiatric disorders. Non-invasive brain stimulation using gamma frequency generated positive treatment effects on overall clinical symptoms and cognitive function in SZ and alleviated depressive symptoms in MDD. Of note, higher stimulation frequency and longer treatment duration increased the therapeutic effects of gamma frequency neuromodulation in SZ. These findings suggest that non-invasive gamma frequency stimulation holds promise for patients with schizophrenic and depressive disorders. However, our review did not indicate a clear therapeutic advantage of gamma frequency stimulation over control conditions in BD or ASD.

Gamma-frequency neuromodulation approaches in SZ, including iTBS, cTBS, and 40 Hz tACS, were associated with clinically meaningful symptom improvement. This pattern is consistent with a recent study (Kishi et al. (2024)) where iTBS targeting the left dorsolateral prefrontal cortex statistically significantly reduced overall symptom severity (standard mean difference, SMD = −0.57), with moderate effects on negative (SMD = −0.89), depressive (SMD = −0.70), and anxious symptoms (SMD = −0.58), but no significant benefit for positive symptoms relative to sham across TBS protocols (Kishi et al., 2024a). Given that iTBS comprised 62% of studies in the present meta-analysis, this concordance is expected. Notably, stimulation frequency (50 Hz) and treatment duration (2–3 weeks) emerged as significant moderators of response, with greater efficacy after longer treatment durations (>2 weeks) suggesting cumulative rather than acute effects. However, these results should be interpreted cautiously, as estimates were driven by a limited number of studies.

Consistent with our hypothesis, gamma frequency neuromodulation was associated with a moderate improvement in overall cognitive performance in schizophrenia. This finding aligns with prior meta-analytic evidence demonstrating significant working memory gains following brain stimulation in schizophrenia (Kishi et al., 2024a; Martin et al., 2017). In contrast, a recent meta-analysis focusing on 40 Hz light-and-sound stimulation reported no significant cognitive benefits but observed large effects on brain structural measures in Alzheimer’s disease (SMD = 1.74) (Ang et al., 2025), suggesting potential disorder-specific mechanisms and outcome sensitivity. Similarly, Begemann et al. found no cognitive improvement following transcranial magnetic stimulation in schizophrenia (Begemann et al., 2020), which may be attributable to the predominance of non-gamma frequencies (e.g., 10–20 Hz) in the included studies. In contrast, gamma-targeted protocols have shown more promising cognitive effects in schizophrenia: iTBS has been associated with working memory enhancement (L. Li et al., 2025), and 40 Hz tACS demonstrated a trend toward improved verbal learning relative to sham (Cao et al., 2025).

Gamma frequency stimulation was also associated with modest improvements in depressive symptoms. Effect sizes were remarkably consistent across diagnoses, with small effects observed in SZ (k = 8, SMD = −0.39), MDD (k = 23, SMD = −0.34), and BD (k = 6, SMD = −0.43), suggesting a broadly comparable but limited antidepressant effect of gamma frequency neuromodulation. This pattern is supported by a meta-analysis of TBS for depression, in which cTBS targeting the right dorsolateral prefrontal cortex (DLPFC) combined with iTBS to the left DLPFC accounted for most observed antidepressant effects (Kishi et al., 2024b). Convergent evidence from open-label and double-blind trials using 50 Hz TBS (Cole et al., 2021, 2020) and 40 Hz violet light sensory stimulation (Noda et al., 2025) further demonstrated significant reductions in depressive symptoms alongside cognitive improvements. In contrast, 40 Hz tACS did not show significant antidepressant effects relative to sham (Alexander et al., 2019b), suggesting that sensory stimulation might be a more effective treatment option at this frequency.

Interestingly, contrary to our initial hypothesis, we did not observe modality-specific differences in clinical efficacy across SZ and MDD, while frequency emerged as a potentially important factor regardless of stimulation modality. Most gamma-frequency interventions seek to induce cortical entrainment via externally imposed rhythmic stimulation, regardless of the delivery method (Hanslmayr et al., 2019). However, it is important to note that the majority of studies included in this review employed magnetic/electrical stimulation-based approaches such as TBS or tACS, whereas only approximately 2% of studies utilized sensory stimulation paradigms. These modalities likely engage neural systems through fundamentally different mechanisms. TBS and tACS deliver focal stimulation to targeted cortical regions, primarily modulating local cortical excitability and network activity. In contrast, rhythmic sensory stimulation (e.g., light and auditory stimulation) can drive large-scale neural entrainment through sensory pathways and thalamocortical circuits, potentially synchronizing activity across widespread brain networks. This large-scale entrainment is a key mechanistic feature of sensory stimulation approaches and may differentiate them from more focal neuromodulation techniques such as TMS. Therefore, directly comparing clinical outcomes across these modalities may be conceptually challenging, as the underlying neural mechanisms and spatial scales of modulation differ substantially. Although clinical benefits have been reported for neuromodulation approaches such as TMS and tACS, effect sizes vary widely across studies and patient populations, precluding firm conclusions regarding comparative efficacy [113–115]. In SZ, findings from 40 Hz tACS (Cao et al., 2025; Wang et al., 2025), iTBS (Chauhan et al., 2021; Vergallito et al., 2024), and cTBS (Chen et al., 2019b; Kang et al., 2024) have been inconsistent, while in MDD the evidence for newer gamma-frequency modalities—including 40 Hz tACS and 40 Hz sensory stimulation—remains sparse, with small effect sizes (SMD = 0.3) and no clear benefit for cTBS.

Current evidence does not indicate a clear therapeutic advantage of gamma frequency stimulation over control conditions in BD or ASD; however, conclusions are limited by the small number of available studies and substantial heterogeneity. In BD, both a systematic review (Mutz, 2023) and randomized controlled trials reported no significant differences between active TBS and sham stimulation (Bulteau et al., 2019; McGirr et al., 2021b). Nevertheless, pooled estimates revealed moderate effect sizes for depressive (g = -0.43, k = 6) and manic symptoms (g = -0.33, k = 1), suggesting that gamma frequency neuromodulation may confer potential clinical benefits that warrant further investigation. In contrast, effect sizes in ASD were uniformly small, indicating limited therapeutic efficacy in the current literature. Although moderate cognitive benefits were observed, interpretation is constrained by the fact that all relevant ASD studies originated from a single laboratory (Fig. S2) and exclusively employed TBS protocols.

Overall, our findings underscore the need for future well-powered trials using different stimulation modalities and examining a broader range of gamma frequencies and treatment durations exceeding two weeks. The need for additional studies employing gamma-rhythmic stimulation to systematically evaluate frequency-specific cognitive effects in schizophrenia, particularly in comparison with non-gamma frequency stimulation protocols. Future studies are encouraged to employ gamma-rhythmic stimulation, particularly tACS and sensory-based paradigms such as GENUS, to disentangle frequency-specific mechanisms and establish their therapeutic relevance across neuropsychiatric disorders.

### Limitations

Certain limitations of this meta-analysis warrant acknowledgment. A key interpretational issue is that the majority of included studies used 50 Hz theta-burst stimulation rather than sustained gamma entrainment paradigms. Although TBS contains a 50 Hz burst structure, its physiological and clinical effects may also depend critically on theta-patterned delivery, cortical target, and cumulative stimulation dose. As such, the present findings may better reflect the efficacy of gamma-burst-patterned neuromodulation, particularly TBS, rather than gamma entrainment per se. Future studies should directly compare TBS, gamma-tACS, and 40 Hz sensory stimulation to determine whether clinical effects are mediated by shared gamma mechanisms or by modality-specific pathways. Secondly, a large proportion of the included studies had small sample sizes and insufficient statistical power, which heightens the likelihood of false-negative results. The relatively small number of eligible studies, together with their modest sample sizes, may therefore constrain the robustness and generalizability of the pooled estimates. Second, substantial heterogeneity was observed across studies, likely reflecting variability in study design, stimulation parameters, target regions, and outcome measures. Although such heterogeneity complicates the interpretation of aggregated effects, sensitivity analyses supported the stability of the findings. Specifically, pooled effect sizes were consistent in both direction and magnitude following the sequential exclusion of individual studies, confirming that the overall results were not dependent on any single study.

### Conclusion

The findings of this systematic review and meta-analysis offer preliminary support for the clinical and cognitive benefits of gamma frequency neuromodulation in the context of neuropsychiatric conditions. Non-invasive gamma frequency stimulation demonstrated potentially beneficial effects on global clinical symptoms and cognitive performance in SZ and was associated with symptom improvement in depression. Importantly, stimulation frequency and treatment duration emerged as positive moderators of treatment response in schizophrenia. These findings support the potential clinical utility of non-invasive rhythmic gamma stimulation in neuropsychiatry, especially for patients with schizophrenic and depressive symptomatology. However, the literature remains methodologically heterogeneous and heavily weighted toward TBS-based protocols, limiting strong conclusions about gamma-specific mechanisms or cross-modality efficacy. Future work should directly compare frequency-specific effects (e.g., 40 vs. 50 Hz) and modality-specific impacts (e.g., TMS vs. tACS vs. GENUS) cross disorder to establish optimal stimulation parameters and advance frequency-guided therapeutic strategies.

## Acknowledgments

We are deeply grateful to Dr Diane Chan, Dr Ute Geigenmuller, and Mr Merric Foley, who provided constructive feedback, and Dr Jung Park, who shared his review table with us to include additional references.

## Contributions of authors

Literature searches, screening, and risk of bias assessments were carried out by MX, AS, RP, and MZ. The literature review and organization of study characteristics were handled by MX and AS. Data extraction, meta-analyses, and manuscript drafting were performed by MX, while conceptual development and refinement involved MX, DM, SN, and JM. AB generated the figures using Biorender. LHT and RF oversaw the revision and refinement of the manuscript.

## Declaration of generative AI and AI-assisted technologies in the manuscript preparation process

During the preparation of this work the author(s) used ChatGPT and Scite.AI in order to correct grammar and check citation. After using this tool/service, the author(s) reviewed and edited the content as needed and take(s) full responsibility for the content of the published article.

## Data Availability Statement

Data and R scripts used for meta-analyses are available at GitHub in the following link: https://github.com/xumei-cloud/gamma_stimulation_psychiatry.git

