## Supplementary_materials for "Gamma Frequency Stimulation Provides Therapeutic Potential in Neuropsychiatry: A Systematic Review and Meta-Analysis"

The search strategy will combine two core concepts:

1. ***Gamma Neuromodulation:***
   “Gamma Neuromodulation” OR “Gamma Stimulation” OR “Gamma Entrainment” OR “40 Hz Stimulation” OR “30 Hz Stimulation” OR “50 Hz Stimulation” OR “30-50 Hz Stimulation” OR “40 Hz Neuromodulation” OR “40 Hz Transcranial Magnetic Stimulation” OR “40 Hz Deep Transcranial Magnetic Stimulation” OR “40 Hz Transcranial Alternating Current Stimulation” OR “40 Hz Audiovisual Entrainment” OR “40 Hz Sensory Stimulation” OR “40 Hz Deep Brain Stimulation” OR “40 Hz Vagus Nerve Stimulation” OR “40 Hz Ultrasound Stimulation” OR “40 Hz Subcutaneous Electrical Stimulation”
2. ***Target Conditions:***
   2.1 **Neurodevelopmental Disorders:** Autism Spectrum Disorder, Attention-Deficit/Hyperactivity Disorder
   2.2 **Schizophrenia Spectrum and Other Psychotic Disorders:** Schizotypal (Personality) Disorder, Delusional Disorder, Brief Psychotic Disorder, Schizophreniform Disorder, Schizophrenia, Schizoaffective Disorder
   2.3 **Bipolar and Related Disorders:** Bipolar I Disorder, Bipolar II Disorder, Cyclothymic Disorder
   2.4 **Depressive Disorders:** Disruptive Mood Dysregulation Disorder, Major Depressive Disorder, Persistent Depressive Disorder, Dysthymia, Premenstrual Dysphoric Disorder

**Table S1**
**Study Characteristics**

| Authors | Country | Active group | Control group | Age | Gender | Education | Type of stimulation | Frequency | Pulses per session | Duration | Clinical Assessment | Cognitive task |
|---|---|---|---|---|---|---|---|---|---|---|---|---|
| **Schizophrenia Spectrum Disorders** | | | | | | | | | | | | |
| Cao et al., 2025 | China | 33 | 17 | Control M= 50.65 (9.98) Active M = 47.30 (11.37) | Control: 11M 6F Active: 18M 15F | – | Gamma tACS | 40 Hz | 24000 | 14 days | PANSS | MCCB |
| Hoy et al., 2016 | Australia | 11 | 11 | Control M= 48.06 (11.38) Active M = 45.38 (12.03) | Control 5M 6F Active 5M 6F | Control 14.73 (2.37) Active 14.73 (2.37) | Gamma tACS | 40 Hz | 48000 | 14 days | PANSS | 2-back |
| Wang et al., 2025 | China | 16 | 16 | Control M= 48.06 (11.38) Active M = 45.38 (12.03) | Control 7M 9F Active 7M 9F | Control 48.06 (11.38) Active 11.06 (2.79) | Gamma tACS | 40 Hz | 6720 | 28 days | PANSS | MoCA |
| Basavaraju et al., 2021 | India, USA | 30 | 30 | Control M = 18-50 years Active M = 18-50 years | – | – | iTBS | 50 Hz | 600 | 5 days | SANS | – |
| Bation et al., 2021 | France | 12 | 10 | Control M = 41.60 (12.63) Active M = 42.33 (9.44) | Control 9M 1F Active 12F | – | iTBS | 50 Hz | 990 | 10 days | SANS | – |
| Bodén et al., 2021 | Sweden | 28 | 28 | Control M = 31.5 (10.4) Active M = 31.3 (9.7) | Control 16M 12F Active 16M 12F | – | iTBS | 50 Hz | 1200 | 20 days | CAINS MADRS | – |

| | | | | | | | | | | | | |
|---|---|---|---|---|---|---|---|---|---|---|---|---|
| Chauhan et al., 2021 | India | 16 | 14 | Control M = 39.35 (8.22) Active M = 41.74 (8.85) | Control 8M 9F Active 7M 12F | – | iTBS | 50 Hz | 1200 | 5 days | PANSS | – |
| Gao et al., 2025 | China | 50 | 50 | Control M = 58.50 (4.74) Active M = 56.24 (6.98) | Control 29M 21F Active 28M 22F | Control 10.08 (2.67) Active 10.72 (2.59) | iTBS | 50 Hz | 600 | 28 days | PANSS SANS | – |
| Jin et al., 2023 | China | 34 | 32 | Control M = 47.47 (10.99) Active M = 47.60 (9.46) | Control 19M 13F Active 18M 16F | Control 7.00 (2.30) Active 7.53 (2.36) | iTBS | 50 Hz | 1800 | 28 days | PANSS | – |
| Kos et al., 2024 | Netherlands | 32 | 16 | Control M = 34.0 Active M = 33.0 | Control 12M 4F Active 24M 8F | Control 16.0 (2.8) Active 17.1 (2.9) | iTBS | 30 Hz | 990 | 1 day | AES PANSS SANS | – |
| Li et al., 2025 | Taiwan | 20 | 19 | Control M = 45.95 (7.97) Active M = 49.15 (5.12) | Control 10M 9F Active 12M 8F | – | iTBS | Hz | 1200 | 20 days | PANSS | MoCA |
| Li et al., 2025 | China | 21 | 20 | Control M = 32.15 (6.23) Active M = 30.29 (7.91) | Control 9M 11F Active 10M 11F | Control 12.25 (2.40) Active 13.52 (2.77) | iTBS | 50 Hz | 600 | 28 days | PANSS | MCCB |
| Li et al., 2024 | China | 33 | 28 | Control M = 30.29 (8.86) Active M = 32.06 (10.59) | Control 16M 12F Active 10M 23F | Control 14.71 (2.79) Active 13.39 (3.27) | iTBS | 50 Hz | 600 | 28 days | PANSS | MCCB |
| Vergallito et al., 2024 | Italy | 7 | 3 | Control M = 32.3 (8.3) Active M = 38.3 (11.2) | Control 1M 4F Active 7M 8F | Control 32.3 (8.3) Active 38.3 (11.2) | iTBS | 50 Hz | 600 | 21 days | PANSS BNSS | MCCB |

| | | | | | | | | | | | | |
|---|---|---|---|---|---|---|---|---|---|---|---|---|
| Walther et al., 2020 | Switzerland | 20 | 20 | Control M = 30.5 (11.5) Active M = 34.3 (12.6) | Control 13M 7F Active 12M 8F | Control 14.8 (2.1) Active 12.2 (2.6) | iTBS | 50 Hz | 600 | 14 days | – | TULIA |
| Wang et al., 2020 | China | 25 | 25 | – | – | – | iTBS | 50 Hz | 600 | 14 days | PANSS SAPS SANS HAMA HAMD | – |
| Wang et al., 2022 | China | 33 | 26 | Control M = 24.152 (4.62) Active M = 23.791 (5.33) | Control 11M 15F Active 15M 18F | Control 11.923(2.189) Active 16.424 (1.521) | iTBS | 50 Hz | 1800 | 14 days | PANSS SAPS SANS HAMA HAMD | MoCA |
| Wu et al., 2021 | China | 16 | 16 | Control M = 26.06 (9.69) Active M = 22.06 (3.33) | Control 13M 7F Active 12M 8F | Control 12.63 (2.41) Active 12.02 (2.49) | iTBS | 50 Hz | 1800 | 14 days | PANSS SAPS SANS HAMA HAMD | – |
| Zhu et al., 2021 | China | 32 | 32 | Control M = 35.34 (6.12) Active M = 35.16 (7.14) | Control 14M 18F Active 18M 14F | Control 10.16 (3.94) Active 10.87 (3.18) | iTBS | 50 Hz | 1800 | 14 days | PANSS | – |
| Chen et al., 2019 | China | 16 | 11 | Control M = 30.91 (11.79) Active M = 26.44 (9.08) | Control 7M 4F Active 10M 6F | Control 11.00(3.13) Active 12.56(2.73) | cTBS | 50 Hz | 1800 | 10 days | AHRS PANSS | – |
| Hua et al., 2024 | China | 32 | 30 | Control M = 27.80 (9.40) Active M = 26.90 (9.20) | Control 15M 15F Active 14M 18F | Control 11.93 (2.63) Active 11.38 (3.39) | rTMS | 50 Hz | 1800 | 14 days | AHRS PANSS HAMA HAMD | – |
| Ji et al., 2023 | China | 26 | 26 | Control M = 26.5 (6.65) Active M = 26.4 (8.63) | Control 13M 13F Active 12M 14F | Control 12.0 (2.74) Active 12.4 (2.89) | cTBS | 50 Hz | 1800 | 15 days | PANSS | – |

| | | | | | | | | | | | | |
|---|---|---|---|---|---|---|---|---|---|---|---|---|
| Kang et al., 2023 | China | 19 | 20 | Control M = 29.88(7.64) Active M = 28.75(9.49) | Control 5M 15F Active 7M 13F | – | cTBS | 50 Hz | 1800 | 10 days | AHRS PANSS HAMA HAMD | – |
| Koops et al., 2016 | Netherlands | 32 | 32 | Control M = 42 (13) Active M = 38 (15) | Control 16M 18F Active 24M 13F | – | cTB-rTMS | 50 Hz | 600 | 5 days | PANSS AHRS | – |
| Tikka et al., 2017 | India | 10 | 10 | Control M = 25.50 (5.00) Active M = 28.40 (2.91) | Control 10M Active 10M | – | cTBS | 50 Hz | 1800 | 14 days | PANSS | Visual Memory Task |
| Tyagi et al., 2022 | India | 25 | 25 | Control M = 33.31 (11.48) Active M = 32.17 (11.22) | Control 17M 12F Active 21M 9F | – | cTBS | 50 Hz | 1200 | 14 days | PANSS | – |
| Shinn et al., 2023 | US | 20 | 20 | sham, active M = 31.9 (7.8) active, sham M = 31.9 (7.8) | sham, active 10M 10F active, sham 10M 10F | – | iTBS | 50 Hz | 1800 | 1 day | – | Interval Discrimination Task |
| **Major Depressive Disorder** | | | | | | | | | | | | |
| Noda et al., 2025 | Japan | 57 | 57 | placebo, active M = 44.9 (10.3) active, placebo M = 44.9 (10.3) | Control 30M 27F Active 30M 27F | placebo, active 15.1 (2.1) active, placebo 15.1 (2.1) | Photobiomodulation therapy (Violet light irradiation) | 40 Hz | 432000 | 28 days | MADRS | – |
| Alexander et al., 2019 | US | 11 | 11 | Control M= 36.69 (13.08) Active M = 36.69 (13.08) | Active (40Hz tACS) 11F Control (10Hz) 11F | – | Gamma tACS | 40 Hz | 97600 | 5 days | MADRS | – |

| | | | | | | | | | | | | |
|---|---|---|---|---|---|---|---|---|---|---|---|---|
| Batail et al., 2023 | US | 12 | 12 | Control M = 50 (17) Active M = 51 (15) | Control 8M 4F Active 7M 5F | – | iTBS | 50 Hz | 1800 | 5 days | MADRS | – |
| Chou et al., 2023 | Taiwan | 17 | 11 | Control M = 43.5 (12.1) Active M = 49.2 (14.9) | Control 5M 6F Active 8M 9F | – | biTBS | 50 Hz | 600 | 22 days | HAMD21 | – |
| Duprat et al., 2016 | Belgium | 50 | 50 | Control M = 42 (12) Active M = 42 (12) | Control 15M 35F Active 15M 35F | – | iTBS | 50 Hz | 32400 | 4 days | BDI | – |
| Fitzgerald et al., 2020 | Australia | 36 | 38 | Control M = 44.7 (12.2) Active M = 44.0 (12.2) | Control 21M 17F Active 17M 19F | – | iTBS | 50 Hz | 63,000 | 28 days | MADRS | – |
| Holczer et al., 2021 | Hungary | 10 | 10 | Control M = 48.68 (12.35) Active M = 51.86 (14.55) | Control 4M 6F Active 1M 9F | – | biTBS | 50 Hz | 600 | 10 days | HDRS | – |
| Huang et al., 2025 | China | 15 | 22 | Control M = 21.68 (5.1) Active M = 19.93 (3.2) | Control 12M 10F Active 5M 10F | – | iTBS | 50 Hz | 600 | 15 days | BSI BDI HAM-A | – |
| Li et al., 2023 | Taiwan | 24 | 24 | Control M = 37.0 (2.7) Active M = 37.0 (3.1) | Control 9M 15F Active 8M 16F | – | piTBS | 50 Hz | 1800 | 10 days | HDRS | – |
| Li et al., 2014 | Taiwan | 15 | 15 | Control M = 46.9 (25–58) Active M = 42.4 (25–61) | Control 4M 11F Active 7M 8F | – | iTBS | 50 Hz | 1800 | 14 days | HDRS | – |
| Liu et al., 2025 | China | 22 | 18 | Control M = 15.39 (1.94) Active M = 15.50 (1.72) | Control 5M 13F Active 4M 18F | – | iTBS | 50 Hz | 1800 | 2 days | MADRS | – |

| | | | | | | | | | | | | |
|---|---|---|---|---|---|---|---|---|---|---|---|---|
| Stöhrmann et al., 2023 | Austria, Hong Kong | 15 | 5 | Control M = 44.4 (13.2) Active M = 36.1 (11.6) | Control 1M 4F Active 7M 8F | – | iTBS | 50 Hz | 1200 | 21 days | HDRS-17 BDI-II IDS-C | – |
| Tavares et al., 2021 | Brazil | 42 | 48 | Control M = 38.0 (10.85) Active M = 40.8 (9.98) | Control 15M 33F Active 11M 31F | Control 15.33 (3.87) Active 14.31 (4.18) | iTBS | 50 Hz | 1800 | 42 days | MADRS HAM-A YMRS | – |
| Wilkening et al., 2022 | Germany | 40 | 41 | sham, active M = 35.65 (13.03) active, sham M = 35.65 (13.03) | sham, active 26M 15F active, sham 21M 19F | sham, active 16.35 (3.44) active, sham 16.35 (3.44) | iTBS | 50 Hz | 1800 | 14 days | MADRS | – |
| Zavorotnyy et al., 2020 | Germany | 72 | 72 | Control M = 18 - 65 yrs Active M = 18 - 65 yrs | – | – | iTBS | 50 Hz | 1200 | 28 days | HDRS BDI | – |
| Zhang et al., 2024 | China | 29 | 30 | Control M = 14.60 (2.85) Active M = 15.83 (2.41) | Control 22M 8F Active 24M 5F | – | iTBS | 50 Hz | 1200 | 21 days | HAMD-17 | – |
| Zhao et al., 2023 | China | 23 | 22 | Control M = 17.27 (2.59) Active M = 17.13 (1.94) | Control 5M 17F Active 3M 20F | – | iTBS | 50 Hz | 1800 | 14 days | BSI HAMD-24 SDS | – |
| Li et al., 2014 | Taiwan | 15 | 15 | Control M = 46.9 (25–58) Active M = 49.2 (27–64) | Control 4M 11F Active 5M 10F | – | cTBS | 50 Hz | 1800 | 14 days | HDRS-17 | – |
| Garcia et al., 2021 | US | 20 | 20 | Control M = 30.3 (4.7) Active M = 30.3 (4.7) | Control 20F Active 20F | – | VNS | 30 Hz | 30000 | 10 days | BDI STAI | – |
| **Bipolar Disorder** | | | | | | | | | | | | |

| | | | | | | | | | | | | |
|---|---|---|---|---|---|---|---|---|---|---|---|---|
| Luo et al., 2024 | China | 22 | 20 | Control M = 15.75 (1.552) Active M = 15.59 (1.764) | Control 2M 18F Active 4M 18F | Control 9.70 (1.625) Active 9.27 (1.638) | iTBS | 50 Hz | 3600 | 21 days | HAMA HAMD HCL-32 | – |
| McGirr et al., 2021 | Canada | 18 | 19 | Control M = 43.00 (14.34) Active M = 44.78 (13.71) | Control 7M 12F Active 7M 11F | Control 14.95 (2.17) Active 15.72 (3.86) | iTBS | 50Hz | 600 | 21 days | MADRS | – |
| Sheline et al., 2024 | US | 12 | 12 | Control M = 43.6 (19.2) Active M = 43.1 (15.2) | Control 6M 6F Active 6M 6F | – | aiTBS | 50 Hz | 90,000 | 5 days | MADRS | – |
| Dellink et al., 2024 | Belgium | 18 | 19 | Control M = 51 (14) Active M = 48 (7) | Control 6M 13F Active 5M 13F | Control 15 (3) Active 14 (2) | cTBS | 50 Hz | 1800 | 4 days | HDRS | – |
| Mallik et al., 2023 | India | 11 | 9 | Control M = 34.63 (10.55) Active M = 43.00 (12.954) | Control 5M 3F Active 5M 6F | – | cTBS | 50 Hz | 1800 | 5 days | HAMD HAMA | – |
| Diederichs et al., 2021 | Canada | 11 | 10 | Control M = 42.20 (13.32) Active M = 45.27 (14.52) | Control 5M 5F Active 6M 5F | Control 16.00 (2.16) Active 16.64 (3.72) | iTBS | 50 Hz | 600 | 10 days | MADRS | – |
| **Autism Spectrum Disorder** | | | | | | | | | | | | |

| | | | | | | | | | | | | |
|---|---|---|---|---|---|---|---|---|---|---|---|---|
| Ni et al., 2021 | Taiwan | 40 | 35 | Control M = 12.5 (2.9) Active M = 13.0 (2.8) | Control 30M 5F Active 38M 2F | – | iTBS | | 38,400 | 42 days | SRS RBS-R | – |
| Ni et al., 2024 | Taiwan, Canada | 13 | 12 | Control M = 13.1 (6.3) Active M = 12.9(6.7) | Control 11M 1F Active 12M 1F | – | iTBS | 50 Hz | 12,000 | 28 days | SRS RBS-R EDI ABAS-II | – |
| Ni et al., 2017 | Taiwan | 19 | 19 | Control M = 20.8 (1.4) Active M = 20.8 (1.4) | Control 14M 5F Active 14M 5F | – | iTBS | 50 Hz | 600 | 7 days | SRS | WCST CCPT |
| Ni et al., 2022 | Taiwan | 13 | 13 | sham, active M = 22.4 (1.4) active, sham M = 23.2 (1.5) | sham, active 7M 1F active, sham 6M 1F | – | iTBS | 50 Hz | 1200 | 5 days | AQ | WCST |
| Ni et al., 2023 | Taiwan | 30 | 30 | Control M = 13.2 (3.2) Active M = 13.1 (3.4) | Control 27M 3F Active 28M 2F | – | cTBS | 50 Hz | 9600 | 8 weeks | SRS RBS-R EDI ABAS-II | – |

*Note: tACS: Transcranial Alternating Current Stimulation; PANSS: Positive and Negative Syndrome Scale; MCCB: MATRICS Consensus Cognitive Battery; MoCA: Montreal Cognitive Assessment Test; iTBS: Intermittent Theta Burst Stimulation; SANS: Scale for the Assessment of Negative Symptoms; CAINS: Clinical Assessment Interview for Negative Symptoms; MADRS: Montgomery-Åsberg Depression Rating Scale; AES: Apathy Evaluation Scale; BNSS: Brief Negative Symptom Scale; TULIA: Test of Upper Limb Apraxia; SAPS: Scale for the Assessment of Positive Symptoms; HAM-A: Hamilton Anxiety Rating Scale; ; HAMD: Hamilton Rating Scale for Depression; cTBS: Continuous Theta Burst Stimulation; VNS: Vagal nerve stimulation AHRS: Auditory Hallucination Rating Scale; BSI: Beck Scale for Suicidal Ideation; BDI: Beck Depression Inventory; HDRS-17: 17-item Hamilton Depression Rating Scale; HAMD21: 21-item Hamilton Rating Scale for Depression; HDRS: Hamilton Depression Rating Scale; IDS-C: Inventory of Depressive Symptomatology – Clinician Rated; YMRS: Young Mania Rating Scale; SDS: Sheehan Disability Scale; STAI: State-Trait Anxiety Inventory; HCL: Hypomania Checklist; SRS: Social Responsiveness Scale; RBS-R: Repetitive Behavior Scale - Revised; EDI: Emotion Dysregulation Inventory; ABAS-II: Adaptive Behavior Assessment System-II; WCST: Wisconsin Card Sorting Test; CCPT: Conners' Continuous Performance Test; AQ: Autism Spectrum Quotient*

**Table S2**

Summary table of subgroup analysis in SZ and MDD

| Subgroups | k | SMD | 95% CI | $I^2$ | *p* |
|---|---|---|---|---|---|
| **SZ** | | | | | |
| *Stimulation modality* | | | | | 0.19 |
| 40 Hz tACS | 2 | 0.01 | [-0.56; 0.58] | 92.9% | |
| cTBS | 6 | -0.63 | [-1.00; -0.25] | 95.3% | |
| iTBS | 13 | -0.46 | [-0.69; -0.22] | 96.2% | |
| *Frequency* | | | | | <0.001*** |
| 30 Hz | 1 | 0.06 | [-0.12; 0.25] | - | |
| 40 Hz | 2 | 0.01 | [-0.56; 0.58] | 92.9% | |
| 50 Hz | 18 | -0.54 | [-0.74; -0.34] | 95.5% | |
| *Duration* | | | | | <0.001*** |
| <1 week | 1 | 0.06 | [-0.12; 0.25] | - | |
| 1-2 week | 4 | -0.23 | [-0.92; 0.46] | 95.1% | |
| 2-3 week | 8 | -0.61 | [-0.83; -0.38] | 93.2% | |
| >=3 week | 8 | -0.50 | [-0.80; -0.20] | 94.7% | |
| *Pulses per session* | | | | | 0.20 |
| 500-1000 | 9 | -0.41 | [-0.75; -0.07] | 97.2% | |
| 1000-1500 | 2 | -0.41 | [-0.81; -0.01] | 89.1% | |
| 1500-2000 | 8 | -0.65 | [-0.92; -0.38] | 93.8% | |
| >=2000 | 2 | 0.01 | [-0.56; 0.58] | 92.9% | |
| **MDD** | | | | | |
| *Stimulation modality* | | | | | 0.56 |
| 40 Hz tACS | 1 | -0.40 | [-0.80; -0.00] | - | |
| BiTBS | 5 | -0.35 | [-0.97; 0.27] | 93.0% | |
| cTBS | 1 | -0.06 | [-0.32; 0.20] | - | |
| iTBS | 14 | -0.37 | [-0.74; -0.01] | 96.8% | |
| SS | 1 | -0.21 | [-0.35; -0.07] | - | |
| VNS | 1 | -0.32 | [-0.53; -0.11] | - | |
| *Frequency* | | | | | |
| 30 Hz | 1 | -0.32 | [-0.53; -0.11] | - | 0.64 |
| 40 Hz | 2 | -0.23 | [-0.36; -0.10] | 0.0% | |
| 50 Hz | 20 | -0.35 | [-0.64; -0.06] | 95.9% | |
| *Duration* | | | | | 0.37 |
| <1 week | 3 | -0.18 | [-0.31; -0.05] | 33.3% | |
| 1-2 week | 2 | -1.16 | [-2.65; 0.34] | 95.7% | |
| 2-3 week | 9 | -0.44 | [-0.86; -0.02] | 95.4% | |
| >=3 week | 9 | -0.12 | [-0.45; 0.21] | 91.0% | |
| *Pulses per session* | | | | | 0.32 |
| 500-1000 | 4 | 0.08 | [-0.43; 0.59] | 92.4% | |
| 1000-1500 | 4 | -0.26 | [-0.94; 0.42] | 87.7% | |
| 1500-2000 | 12 | -0.52 | [-0.90; -0.14] | 96.2% | |
| >=2000 | 3 | -0.26 | [-0.37; -0.14] | 0.0% | |

*Note:* SZ: Schizophrenia; MDD: Mild Depressive Disorder; k: the number of studies; SMD: standardised mean difference; CI: confidence interval; $I^2$: total heterogeneity; **p* < 0.5; ***p* < 0.01; ****p* < 0.001.

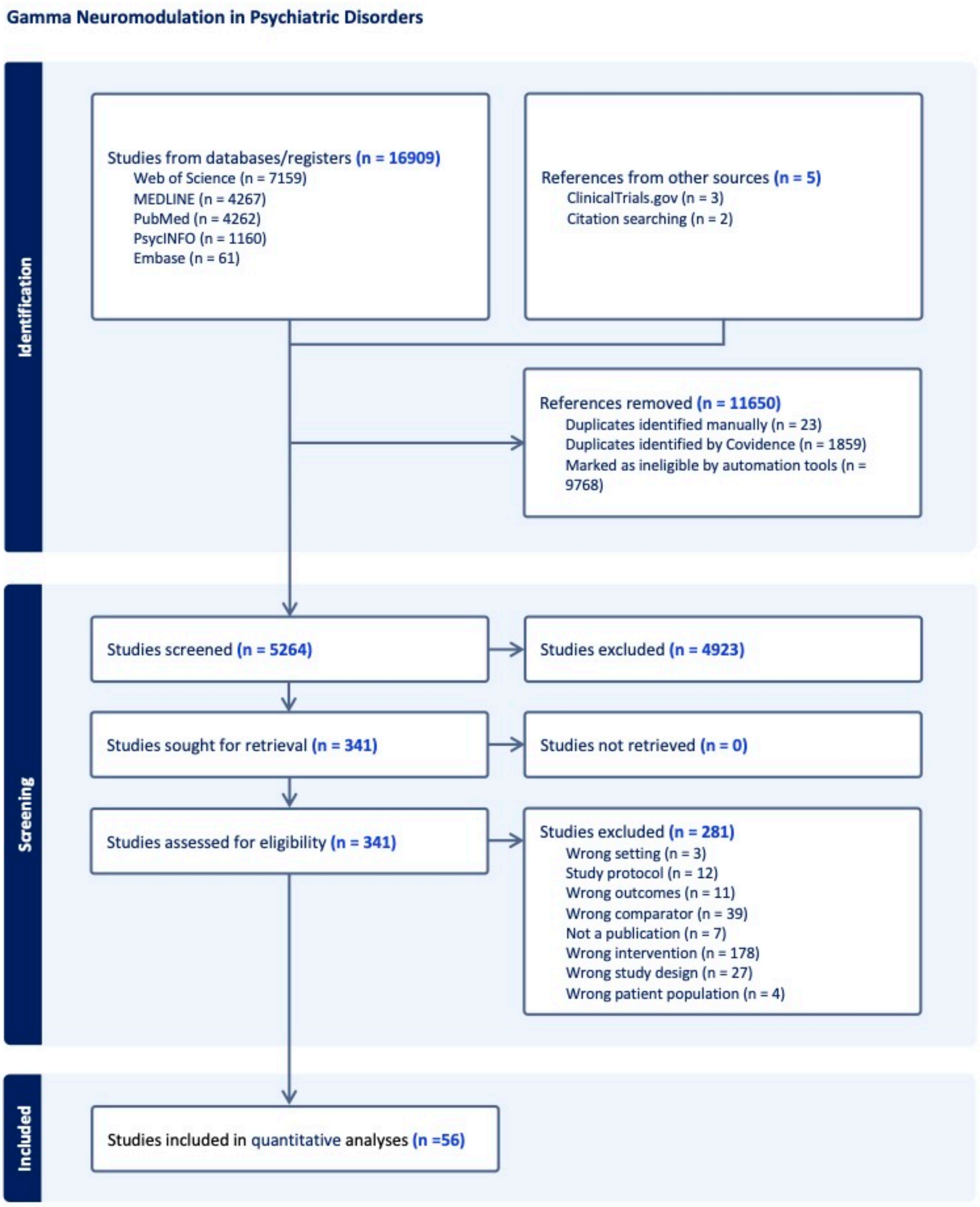


**Fig. S1** Study search and selection workflow

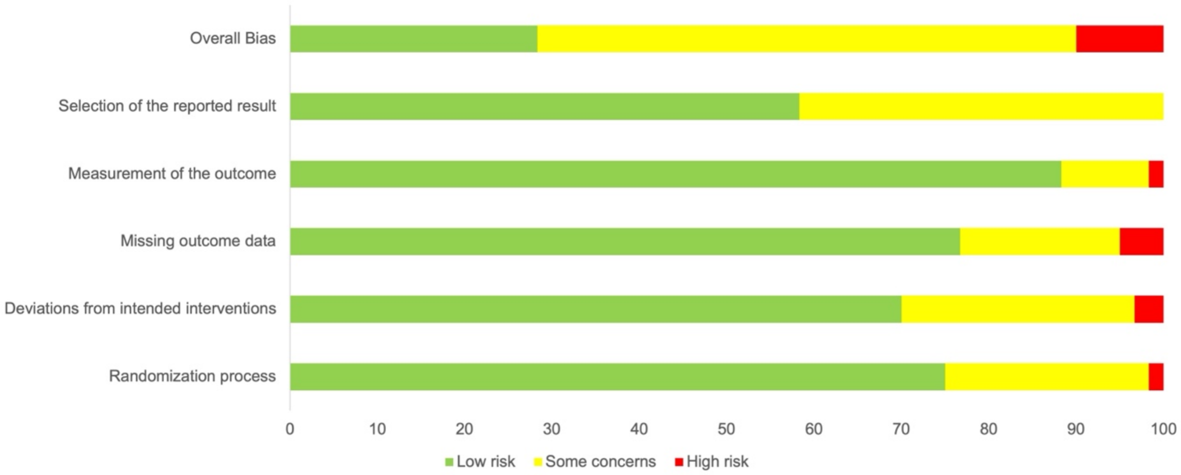


**Fig. S2** Risk of Bias

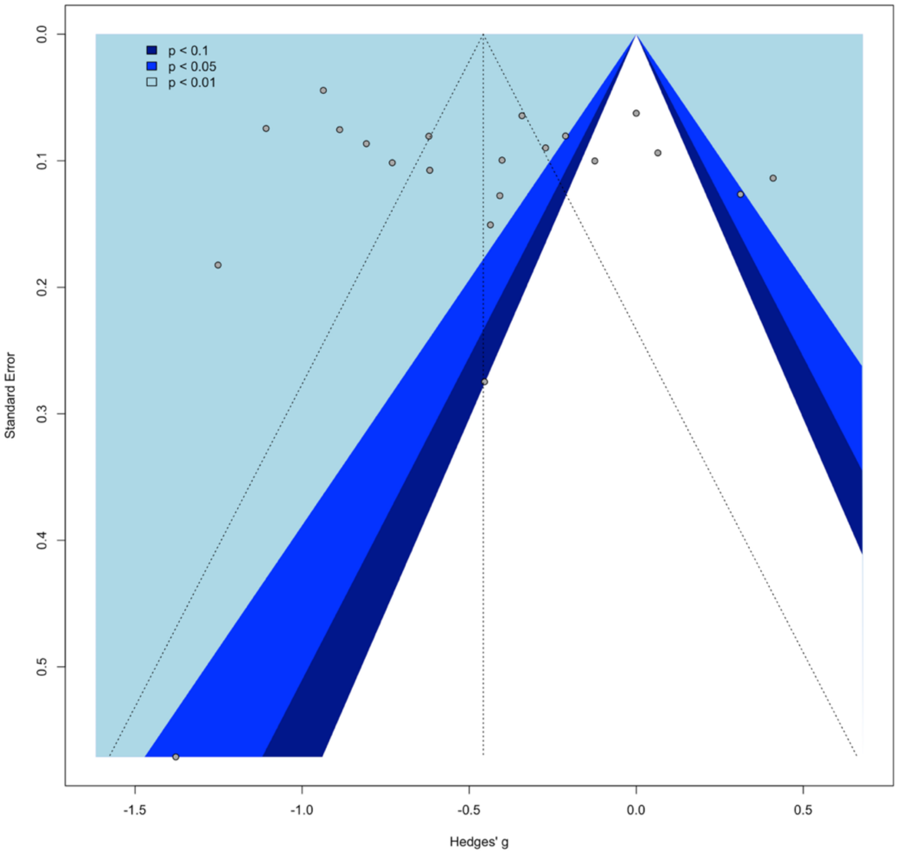


**Fig.S3** Funnel plot of overall clinical symptoms in SZ

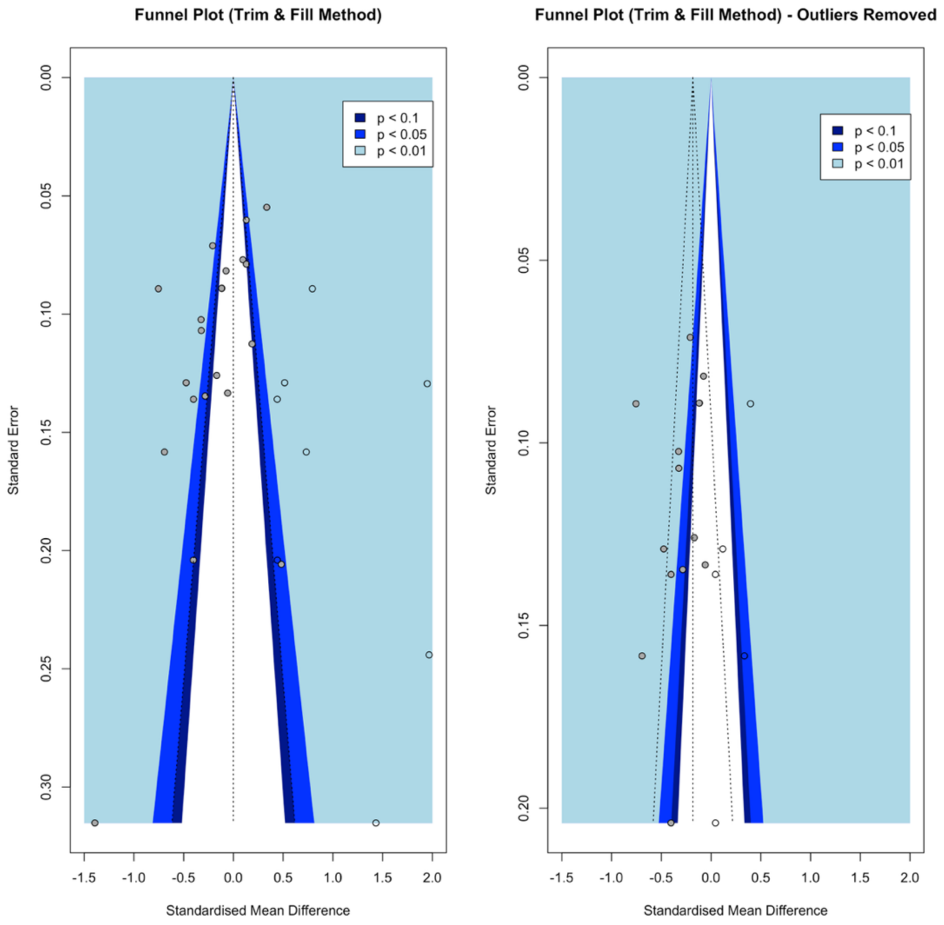


**Fig. S4** Funnel plots in MDD with trim and fill methods

*Notes:* The imputed studies are represented by circles that have no fill color. Outliers include Batail 2023, Fitzgerald 2020, Huang 2025, Stohrmann 2023, Tavares 2021, Tavares 2021, Zhao 2023, Holczer 2021, Zhang 2024a.

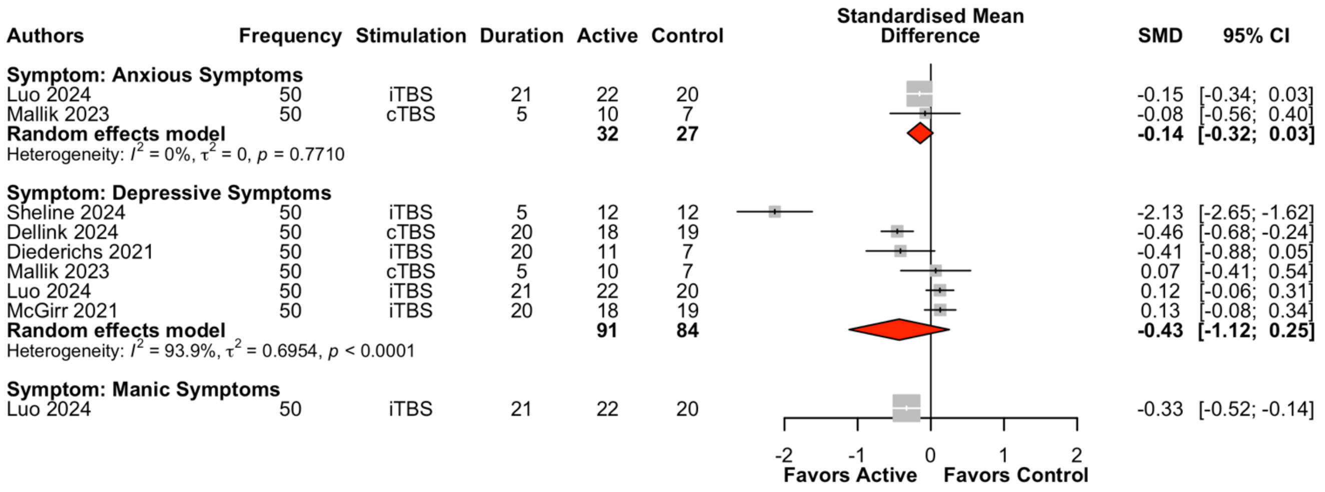


**Fig. S5** Forest plot of gamma stimulation in BD

*Note:* Duration: treatment period (days). iTBS: Intermittent Theta Burst Stimulation; cTBS: Continuous Theta Burst Stimulation; SMD: Standardised Mean Difference; $I^2$: total heterogeneity

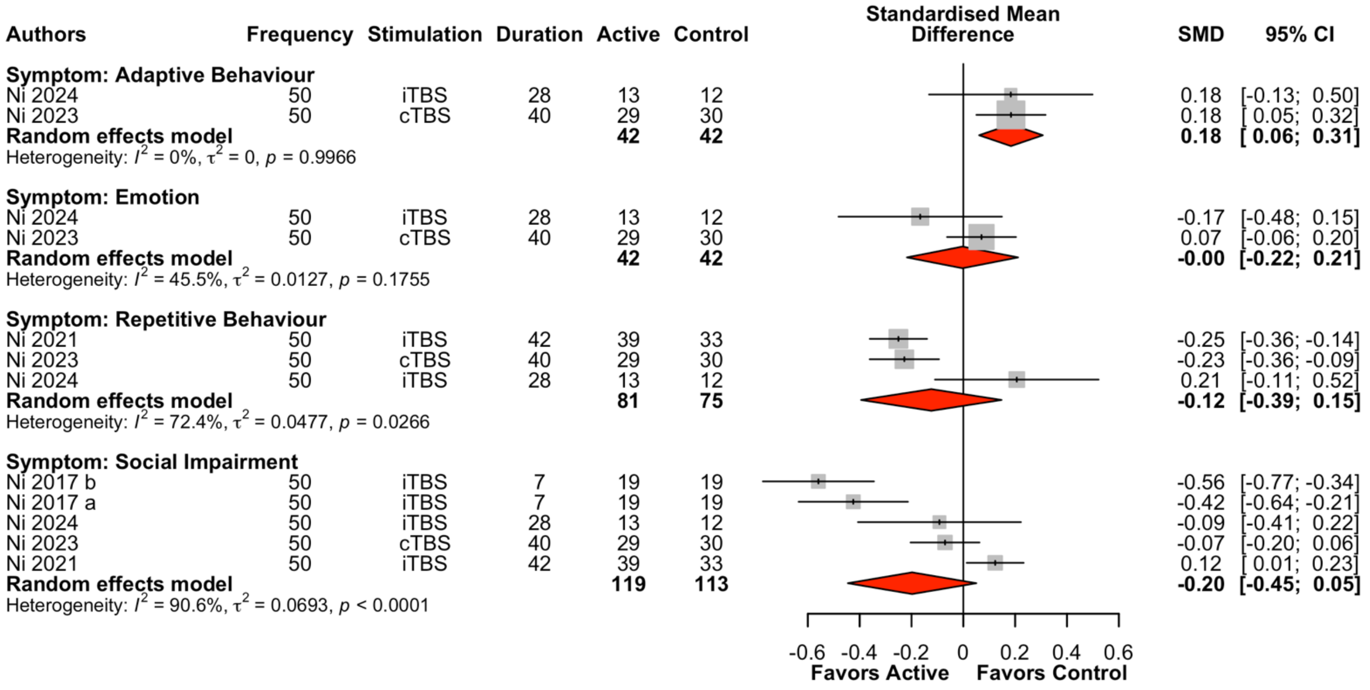


**Fig. S6** Forest plot of gamma stimulation for clinical symptoms in ASD

*Note:* Duration: treatment period (days). iTBS: Intermittent Theta Burst Stimulation; cTBS: Continuous Theta Burst Stimulation; SMD: Standardised Mean Difference; $I^2$: total heterogeneity

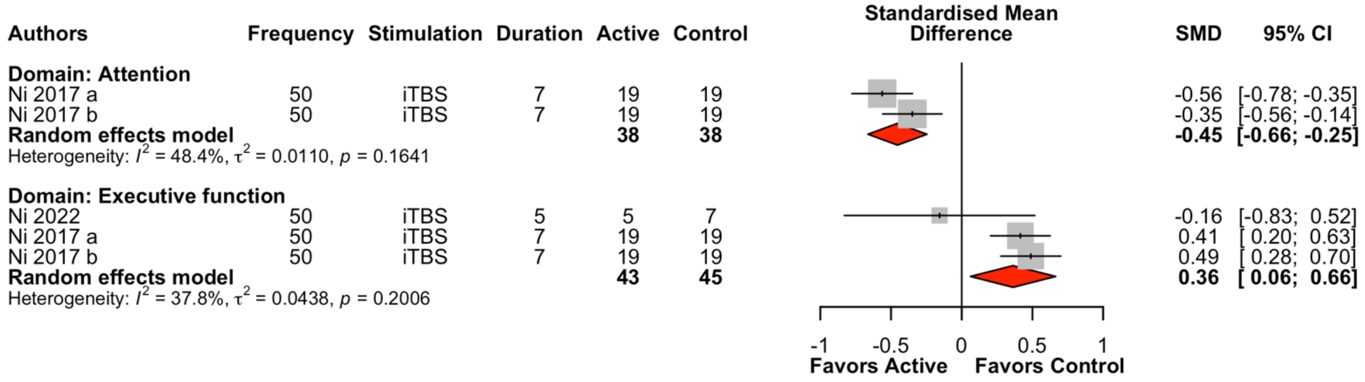


**Fig. S7** Forest plot of gamma stimulation for cognitive functions in ASD

*Note:* Duration: treatment period (days). iTBS: Intermittent Theta Burst Stimulation; SMD: Standardised Mean Difference; $I^2$: total heterogeneity
